# Multi-ancestry transcriptome-wide association studies of cognitive function, white matter hyperintensity, and Alzheimer’s disease

**DOI:** 10.1101/2024.01.03.24300768

**Authors:** Dima L. Chaar, Zheng Li, Lulu Shang, Scott M. Ratliff, Thomas H. Mosley, Sharon L.R. Kardia, Wei Zhao, X. Zhou, J.A. Smith

## Abstract

Genetic variants increase the risk of neurocognitive disorders in later life including Vascular Dementia (VaD) and Alzheimer’s disease (AD), but the precise relationships between genetic risk factors and underlying disease etiology are not well understood. Transcriptome-wide association studies (TWAS) can be leveraged to better characterize the genes and biological pathways underlying genetic influences on disease. To date, almost all existing TWAS have been conducted using expression studies from individuals of a single genetic ancestry, primarily European. Using the joint likelihood-based inference framework in Multi-ancEstry TRanscriptOme-wide analysis (METRO), we leveraged gene expression data from European (EA) and African ancestries (AA) to identify genes associated with general cognitive function, white matter hyperintensity (WMH), and AD. Regions were fine-mapped using Fine-mapping Of CaUsal gene Sets (FOCUS). We identified 266, 23, 69, and 2 genes associated with general cognitive function, WMH, AD (using EA GWAS summary statistics), and AD (using AA GWAS), respectively (Bonferroni-corrected alpha=P<2.9−10^-6^), some of which were previously identified. Enrichment analysis showed that many of the identified genes were in pathways related to innate immunity, vascular dysfunction, and neuroinflammation. Further, downregulation of *ICA1L* was associated with higher WMH and with AD, indicating its potential contribution to overlapping AD and VaD neuropathology. To our knowledge, our study is the first TWAS of cognitive function and neurocognitive disorders that used expression mapping studies in multiple ancestries. This work may expand the benefits of TWAS studies beyond a single ancestry group and help to identify gene targets for pharmaceutical or preventative treatment for dementia.

**Author Summary:** Transcriptome-wide association studies (TWAS) can be used to understand the mechanisms of gene expression that underly disease etiology. However, to date, TWAS methods have mostly been used in a single ancestry group, especially European ancestry (EA), and few TWAS have focused on cognitive function or structural brain measures. We used a newly developed TWAS method called the Multi-ancEstry TRanscriptOme-wide analysis (METRO) to incorproate gene expression data from 801 EA and 1,032 African ancestry (AA) adults to identify genes associated with general cognitive function, structural brain changes called white matter hyperintensities (WMH) that predispose people to vascular dementia, and another form of dementia called Alzheimer’s disease (AD). We found that reduced gene expression of *ICA1L* was associated with more WMH and with AD, indicating its potential contribution to overlapping AD and vascular dementia neuropathologies. To our knowledge, our study is the first TWAS of cognitive function and neurocognitive disorders using multiple ancestries. This work may expand the benefits of TWAS studies beyond a single ancestry group and help to identify gene targets for pharmaceutical or preventative treatment for dementia.

## Introduction

Adult-onset dementia is comprised of a group of aging-related neurocognitive disorders caused by the gradual degeneration of neurons and the loss of brain function. These changes lead to a decline in cognitive abilities and impairment of daily activities and independent function. In the United States, Alzheimer’s disease (AD), the most common cause of dementia, affects 6.8 million adults age 65 and older (1). The second most common form of dementia is vascular dementia (VaD), which often co-occurs with AD and is underdiagnosed (1,2). VaD is often difficult to distinguish from AD because these diseases share cognitive symptoms including noticeable impairment in episodic and semantic memory. While AD and VaD often co-occur, each form of dementia has differing pathophysiology that may precede the illness decades prior.

AD is characterized by aggregation of amyloid-beta protein and neurofibrillary tangles in brain tissue (3,4), while VaD may be caused by reduced blood flow to the brain as a result of small vessel disease (SVD) or stroke and is commonly seen in people with hypertension (5). AD is diagnosed based on a battery of memory tests, brain-imaging tests for degeneration of brain cells and laboratory tests to assess the presence of amyloid and tau proteins in cerebrospinal fluid (6). SVD is primarily detected on magnetic resonance imaging (MRI) as white matter hyperintensities (WMH). It has been hypothesized that vascular and neurodegenerative changes in the brain may interact in ways that increase the likelihood of cognitive impairment. A further challenge in the field is distinguishing between individuals who are aging normally from those with dementia pathology.

A greater understanding of the pathological processes that influence cognitive function in older adults is critical for early intervention during the long preclinical or prodromal phase prior to dementia onset, especially in vulnerable populations (7,8). For example, individuals of African ancestry (AA) have a greater burden of and risk for developing dementia compared to Non-Hispanic Whites (9–12). Differences in gene expression, which are influenced by both genetic and non-genetic factors, likely play a role in shaping racial/ethnic health disparities in neurological outcomes. However, the underlying molecular and environmental mechanisms that influence gene expression are not fully understood, especially in populations with non-European ancestries. Given the multifactorial and complex nature of dementia, multi-omic data integration across ancestry groups may lend insight into these disparities, allowing the identification of targets for intervention and treatment in populations that are most at risk (13).

Genome-wide association studies (GWAS) have identified genetic variants associated with cognitive function and dementia; however, most GWAS variants are located in non-coding regions so their functional consequences are difficult to characterize (14). Transcriptome-wide association studies (TWAS) utilize gene expression and genetic data to increase power for identifying gene-trait associations and characterizing transcriptomic mechanisms underlying complex diseases. To date, however, few TWAS have been conducted on cognitive or structural brain measures. Further, previous TWAS have primarily been conducted in populations of European ancestry (EA), but these results cannot always be generalized to other genetic ancestries due to differences in allele frequencies, patterns of linkage disequilibrium (LD), and relationships between SNPs and gene expression between populations (15–18). To better identify gene-trait associations in non-EA ancestries, it is necessary to incorporate results from recent expression quantitative trait locus (eQTL) mapping studies, which identify genetic variants that explain variations in gene expression levels, conducted in different ancestry groups.(19)

Multi-ancEstry TRanscriptOme-wide analysis (METRO)(20) is a TWAS method that uses a joint likelihood-based inference framework to borrow complementary information across multiple ancestries to increase TWAS power. In this study, we used genotype and gene expression data from 1,032 AA and 801 EA from the Genetic Epidemiology Network of Arteriopathy (GENOA) and summary statistics from published GWAS (21–24) to identify genes associated with general cognitive function, white matter hyperintensity, and AD. We then examined the contribution of different ancestry-dependent transcriptomic profiles on the genetrait associations. Greater knowledge of the underlying molecular mechanisms of dementia that are generalizable to both EA and AA is a critical step in evaluating potential causal variants and genes that could be targeted for pharmaceutical development.

## 2. Materials and Methods

### 2.1. Sample

#### The Genetic Epidemiology Network of Arteriopathy (GENOA)

The GENOA study is a community-based longitudinal study aimed at examining the genetic effects of hypertension and related target organ damage (25). EA and AA hypertensive sibships were recruited if at least 2 siblings were clinically diagnosed with hypertension before age 60. All other siblings were invited to participate, regardless of their hypertension status. Exclusion criteria included secondary hypertension, alcoholism or drug abuse, pregnancy, insulin-dependent diabetes mellitus, active malignancy, or serum creatinine levels >2.5mg/dL. In Phase I (1996–2001), 1,854 AA participants (Jackson, MS) and 1,583 EA participants (Rochester, MN) were recruited (25). In Phase II (2000-2004), 1,482 AA and 1,239 EA participants were successfully followed up, and their potential target organ damage from hypertension was measured. Demographics, medical history, clinical characteristics, information on medication use, and blood samples were collected in each phase. After data cleaning and quality control, a total of 1,032 AA and 801 EA with genotype and gene expression data were available for analysis. Written informed consent was obtained from all participants, and approval was granted by participating institutional review boards (University of Michigan, University of Mississippi Medical Center, and Mayo Clinic).

### 2.2. Measures

#### 2.2.1. Genetic Data

AA and EA blood samples were genotyped using the Affymetrix® Genome-Wide Human SNP Array 6.0 or the Illumina 1M Duo. We followed the procedures outlined by Shang et al.(18) for data processing. For each platform, samples and SNPs with a call rate <95%, samples with mismatched sex, and duplicate samples were excluded. After removing outliers identified from genetic principal component analysis, there were 1,599 AA and 1,464 EA with available genotype data. Imputation was performed using the Segmented HAPlotype Estimation & Imputation Tool (SHAPEIT) v.2.r(26) and IMPUTE v.2(27) using the 1000 Genomes project phase I integrated variant set release (v.3) in NCBI build 37 (hg19) coordinates (released in March 2012). Imputation for each genotyping platform was performed separately and then combined. The final set of genotype data included 30,022,375 and 26,079,446 genetic variants for AA and EA, respectively. After removing genetic variants with MAF ≤ 0.01, imputation quality score (INFO score) ≤ 0.4 in any platform-based imputation, and indels, a total of 13,793,193 SNPs in AA and 7,727,215 SNPs in EA were available for analysis. We used the GENESIS package(28) in R to infer population structure in the analytic sample, and the PC-AiR function was used to extract the first five genotype PCs which were subsequently used to adjust for population structure.

#### 2.2.2. Gene Expression Data

Gene expression levels were measured from Epstein-Barr virus (EBV) transformed B-lymphoblastoid cell lines (LCLs) created from blood samples from a subset of GENOA AA (n=1,233) and EA (n=919). Gene expression levels of AA samples were measured using the Affymetrix Human Transcriptome Array 2.0, while gene expression levels of EA samples were measured using Affymetrix Human Exon 1.0 ST Array. We followed the procedures outlined by Shang et al.(18) In particular, the Affymetrix Expression Console was used for quality control and all array images passed visual inspection. In AA, 28 samples were removed due to either low signal-to-noise ratio (n=1), abnormal polyadenylated RNA spike-in controls (Lys < Phe < Thr < Dap; n=24), sample mislabeling (n=2), or low RNA integrity (n=1), resulting in a total of n=1,205 AA samples for analysis. In EA, duplicated samples (n=31), control samples (n=11) and sex mismatch samples (n=2) were removed, resulting in n=875 EA samples for analysis. We processed data in each population separately. Raw intensity data were processed using the Affymetrix Power Tool software (29). AffymetrixCEL files were normalized using the Robust Multichip Average (RMA) algorithm which included background correction, quantile normalization, log_2_-transformation, and probe set summarization.(30) The algorithm also includes GC correction (GCCN), signal space transformation (SST), and gain lock (value=0.75) to maintain linearity. The Brainarray custom CDF(31) v.19 was used to map the probes to genes. This custom CDF uses updated genomic annotations and multiple filtering steps to ensure that the probes used are specific for the intended gene cluster. Specifically, it removes probes with non-unique matching cDNA/EST sequences that can be assigned to more than one gene cluster. As a result, gene expression data processed using custom CDF are expected to be largely free of mappability issues. After mapping, ComBat(32) was used to remove batch effects. For each gene, we applied a linear regression model to adjust for age, sex, and first five genotype principal components (PCs). We then extracted the residuals and quantile normalized residuals across all samples. We analyzed a common set of 17,238 protein coding genes that were annotated in GENCODE (release 12) (33).

#### 2.2.3. GWAS summary statistics

We used summary statistics from GWAS for general cognitive function (21), WMH (22), AD in EA(23), and AD in AA(24) as input for METRO. Three of the GWASs, Davies et al. (2018), Sargurupremraj et al. (2020), and Bellenguez et al. (2022), were selected because they are the largest meta-analyses to date with publicly available summary statistics; however, we note that all three were conducted in primarily EA samples. We also selected the Kunkle et al. (2021) GWAS because it is the largest meta-analysis to date with public available summary statistics in primarily AA samples. Below, we describe each GWAS and also provide information about the corresponding TWAS analyses that were reported in two of the input GWAS (WMH(22) and AD in EA(23)) which use the same GWAS summary statistics as our analysis but different gene expression data.

##### General cognitive function

We obtained GWAS summary statistics for general cognitive function from a meta-analysis by Davies et al. (2018) that includes the Cohorts for Heart and Aging Research in Genomic Epidemiology (CHARGE), the Cognitive Genomics Consortium (COGENT) consortia and the UK Biobank (UKB; Table 1) (21). This study included 300,486 EA individuals with ages between 16 and 102 years from 57 population-based cohorts. This is the largest available GWAS for general cognitive function, and there are currently no large-scale GWAS studies available in non-EA. General cognitive function was constructed from a number of cognitive tasks. Each cohort was required to have tasks that tested at least three different cognitive domains. Principal component (PC) analysis was performed on the cognitive tests scores within each cohort, and the first unrotated component was extracted as the measure of general cognitive function. Models performed within each cohort were adjusted for age, sex, and population stratification. Exclusion criteria included clinical stroke (including self-reported stroke) or prevalent dementia.

**Table 1.** Sample characteristics of expression quantitative trait locus (eQTL) mapping study and genome-wide association studies (GWAS) participants.

| *eQTL mapping study: Genetic Epidemiology Network of Arteriopathy (GENOA)* |  | Mean (SD) or N (%) or N |
| --- | --- | --- |
| N |  | N=1833 |
| Age (years) |  | 56.85 (10.0) |
| Female |  | 1202 (65.6%) |
| Race/Ethnicity |  |  |
| African Americans |  | 1032 (56.3%) |
| European Americans |  | 801 (43.7%) |
| *General cognitive function GWAS: CHARGE, COGENT, UKB<sup>a</sup> (Davies et al., 2018)* |  | Mean (SD) or N (%) or N |
| N |  | 300,486 |
| Age (years) |  | 56.91 (7.8) |
| Female |  | 52.20% |
| Excluded for dementia and/or stroke diagnosis |  | N=4919 |
| *White matter hyperintensity (WMH) GWAS: CHARGE and UKB<sup>a</sup> (Sargurupremraj et al., 2020)* |  | Mean (SD) or N (%) or N |
| N |  | 48,454 |
| Age (years) |  | 64.17 |
| Female |  | 29215 (57.6%) |
| WMH volume (cm<sup>3</sup>) |  | 7.06 (8.8) |
| Excluded for stroke or pathologies |  | N=1572 |
| *EA Alzheimer's Disease (AD) GWAS: EADB, GR@ACE, EADI, GERAD/PERADES, DemGene, Bonn, the Rotterdam study, the CCHS study, NxC and the UKB<sup>a</sup> (Bellenguez et al., 2022)* |  | Mean (SD) or N (%) or N |
| Discovery sample |  |  |
| AD cases |  | N=39,106 |
| AD proxy cases |  | N=46,828 |
| Controls |  | N=401,577 |
| Age (years) |  |  |
| AD cases or proxy cases |  | 73.55 (8.1) |
| controls |  | 67.86 (8.6) |
| Female |  |  |
| AD cases or proxy cases |  | 62.90% |
| controls |  | 56.10% |
| *AA Alzheimer's Disease (AD) GWAS: AD Genetics Consortium<sup>b</sup> (Kunkle et al., 2021)* |  | Mean (SD) or N (%) or N |
| N |  | 7,970 |
| AD cases |  | 2,748 (34.5%) |
| controls |  | 5,222 (65.5%) |
| Age (years) |  | 74.2 (13.6) |
| Female |  |  |
| AD cases |  | 1,944 (69.8%) |
| controls |  | 3,743 (71.7%) |

Abbreviations: EA, European ancestry; AA, African ancestry
<sup>a</sup> GWAS include only European ancestry participants.<sup>b</sup> GWAS includes only African ancestry participants.

##### White matter hyperintensity

We obtained the GWAS summary statistics for WMH from a meta-analysis conducted by Sargurupremraj et al. (2020) that included 48,454 EA and 2,516 AA with mean age of 66.0 (SD=7.5) years from 23 population-based studies from the CHARGE consortium and UKB (Table 1) (22). We obtained publicly available GWAS summary statistics from only EA individuals. Summary statistics for only EA are publicly available for this GWAS. WMH was measured from MRI scans obtained from scanners with field strengths ranging from 1.5 to 3.0 Tesla and interpreted using a standardized protocol blinded to clinical or demographic features. In addition to T1 and T2 weighted scans, some cohorts included fluid-attenuated inversion recovery (FLAIR) and/or proton density (PD) sequences to measure WMH from cerebrospinal fluid. WMH volume measures were inverse normal transformed, and models adjusted for sex, age, genetic PCs and intracranial volume (ICV). Exclusion criteria included history of stroke or other pathologies that influence measurement of WMH at the time of MRI.

To functionally characterize and prioritize individual WMH genomic risk loci, Sargurupremraj et al.(22) (2020) conducted TWAS using TWAS-Fusion(34) with summary statistics from the WMH SNP-main effects (EA only) analysis and weights from gene expression reference panels from blood (Netherlands Twin Registry; Young Finns Study), arterial (Genotype-Tissue Expression, GTEx), brain (GTEx, CommonMind Consortium) and peripheral nerve tissue (GTEx). This study did not perform fine-mapping following TWAS analysis.

##### Alzheimer’s disease (GWAS in EA)

We obtained the EA GWAS summary statistics for Alzheimer’s disease from stage I meta-analysis by Bellenguez et al. (2022) that included EA from the European Alzheimer and Dementia Biobank (EADB), GR@ACE, EADI, GERAD/PERADES, DemGene, Bonn, the Rotterdam study, CCHS study, NxC and the UKB (Table 1) (23). The meta-analysis was performed on 39,106 clinically diagnosed AD cases, 46,828 proxy-AD and related dementia (ADD) cases, and 401,577 controls. AD cases were clinically diagnosed in all cohorts except UKB, where individuals were identified as proxy-ADD cases if their parents had dementia. Participants without the clinical diagnosis of AD, or those without any family history of dementia, were used as controls. Models performed within each cohort were adjusted for PCs and genotyping centers, when necessary.

To examine the downstream effects of new AD-associated variants on molecular phenotypes in various AD-relevant tissues, Bellenguez et al. (2022) conducted a TWAS with stage I AD GWAS results. The TWAS was performed by training functional expression and splicing reference panels based on the Accelerating Medicines Partnership (AMP)-AD bulk brain and EADB lymphoblastoid cell lines (LCL) cohorts, while leveraging pre-calculated reference panel weights(35) for the GTEx dataset(36) in tissues and cells of interest. TWAS associations were then fine-mapped using Fine-mapping Of CaUsal gene Sets (FOCUS) (37).

##### Alzheimer’s disease (GWAS in AA)

We also acquired the AA GWAS summary statistics for Alzheimer’s disease from meta-analysis by Kunkle et al. (2021)(24) that included individuals of African American ancestry from 15 cohort studies from the AD Genetics Consortium (ADGC; Table 1). The meta-analysis was performed on 2,748 clinically diagnosed AD cases and 5,222 controls with a mean age of 74.2 years (SD=13.6). Models performed within each cohort were adjusted for age, sex, and PCs for population substructure.

### 2.3 Statistical Methods

#### 2.3.1. Multi-ancestry transcriptome-wide association study

Using the Multi-ancEstry TRanscriptOme-wide analysis (METRO) (20), we conducted a high-powered TWAS with calibrated type I error control to identify the key gene-trait associations and transcriptomic mechanisms underlying general cognitive function, WMH and AD. Since gene expression prediction models constructed in different ancestries may contain complementary information, even when the input GWAS was conducted in a single ancestry (20), we used METRO to model gene expression from EA and AA simultaneously. METRO uses a joint-likelihood framework that accounts for SNP effect size heterogeneity and LD differences across ancestries. The framework selectively upweights information from the ancestry that has greater certainty in the gene expression prediction model, increasing power and allowing characterization of the relative contribution of each ancestry to the TWAS results.

METRO is described in Li et al.(20) Briefly, each gene is examined separately using gene expression data from *M* different genetic ancestries. ***Z_m_***is the *n_m_*-vector of gene expression measurements on *n_m_*individuals in the *m*^th^ ancestry with *m*∈{1,…,M}. For the gene of interest, all *cis*-SNPs (*p*), which are in potential linkage disequilibrium (LD) with each other, were extracted as predictors for gene expression. ***G_m_*** is denoted as the *n_m_ x p* genotype matrix for these *cis*-SNPs. Besides the gene expression data, we also used GWAS summary statistics from *n* individuals for an outcome trait of interest. *γ* is the *n*-vector of outcome measurements in the GWAS data and ***G*** is the corresponding *n x p* genotype matrix on the same set of *p cis*-SNPs. The expression vector ***z_m_***, the outcome vector *γ,* and each column of the genotype matrixes are centered and standardized. ***G_m_*** and ***G*** have a mean of zero and variance of one. For each TWAS, we used GWAS summary statistics in the form of marginal z-scores and a SNP-SNP correlation (LD) matrix estimated with genotype data from our GENOA sample that correspond with the ancestry of the GWAS (EA or AA). The following equations describe the relationships between the SNPs, gene expression and the outcome:

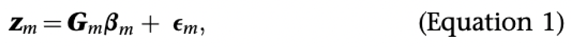

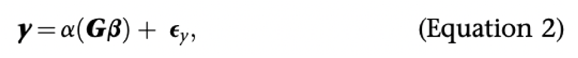

Equation (1) describes the relationship between gene expression and the *cis-*SNP genotypes in the gene expression study in GENOA for the *m*^th^ ancestry (EA or AA). *β_m_* is a *p* vector of the *cis*-SNP effects on the gene expression in the *m^t^*^h^ ancestry and *ε_m_* is an *n_m_*-vector of residual errors with each element following an independent and normal distribution N(0, *σ^2^_m_*) with an ancestry specific variance *σ^2^_m_*. Equation (2) describes the relationship between the genetically regulated gene expression (GReX), calculated from estimated SNP prediction weights, and the outcome trait (general cognitive function, WMH or AD) from the GWAS. There, *Gβ* denotes an *n*-vector of GReX constructed for the GWAS individuals, where *β* = Σ_m_ w_m_*β_m_* is a *p*-vector of SNP effects on the gene expression in the GWAS data, where the weights Σ^M^_m=1_ *w_m_*=1 and *w_m_*≥0. The alpha value (*α*) is the effect of GReX constructed for the GWAS individuals on the outcome trait, and *ε_y_* is an *n_m_*-vector of residual errors with each element following an independent and normal distribution N(0, *σ^2^*). Both equations, specified based on separate studies, are connected through the predictive SNP effects on the gene expression (*β_m_* and *β*). A key assumption made is that the SNP effects on the gene expression in the GWAS, *β*, can be expressed as a weighted summation of the SNP effects on gene expression in the expression studies conducted across ancestries.

We derived the overall GReX effect *α* and the contribution weight of each ancestry (w_1_ for AA and w_2_ for EA) to infer the extent and contribution of the two genetic ancestries in informing the GReX-trait association. The joint model defined in Equations 1 and 2 allows us to borrow association strength across multiple ancestries to enable powerful inference of GReX-trait associations for general cognitive function, WMH and AD. We declared the gene to be significant if the p-value was below the corresponding Bonferroni corrected threshold for the number of tested genes (P<0.05/17,238 = 2.90−10^-6^). Manhattan plots and quantile-quantile (QQ) plots were generated using the *qqman*(38) R package.

#### 2.3.2 Fine-mapping analysis

Since genes residing in the same genomic region may share eQTLs or contain eQTL SNPs in LD with each other, TWAS test statistics for genes in the same region can be highly correlated, making it difficult to identify the true biologically relevant genes among them. To prioritize the putatively causal genes identified by METRO for general cognitive function, WMH, and AD, we conducted TWAS fine-mapping using FOCUS (Fine-mapping Of CaUsal gene Sets) (37). To identify a genomic region with at least one significant gene detected by METRO, we obtained a set of independent, non-overlapping genomic regions, or LD blocks, using Ldetect (39). In each analyzed genomic block, using a standard Bayesian approach, we assigned a posterior inclusion probability (PIP) for each gene to be causal, given the observed TWAS statistics. We used gene-level Z scores, created from p-values using the inverse cumulative distribution function (CDF) of a standard normal distribution, as input into FOCUS. We then ranked the PIPs and computed the 90%-credible set that contains the causal gene with 90% probability. In the FOCUS analysis, a null model which assumes none of the genes in the region are causally associated with the trait is also considered as a possible outcome and may be included in the credible set. Through fine-mapping, we narrowed down significantly associated genes identified by METRO to a shorter list of putatively true associations.

#### 2.3.3 Characterization of identified genes

To interpret our TWAS findings, both before and after fine-mapping, we further examined whether the genes identified by METRO overlapped with those previously identified by their corresponding input GWAS. We created a set of Venn diagrams of overlapping genes identified using METRO with those from the SNP-based GWAS association results(21–24) mapped to the nearest gene using the *VennDiagram* R package (40). We then constructed a second set of Venn diagrams showing overlapping genes identified using METRO with genes identified by gene-based association analyses in each of the input GWAS studies. The gene-based analyses were conducted using MAGMA(41) (general cognitive function,(21) WMH(22), and AD (AA GWAS)(24)) or gene prioritization tests (AD (EA GWAS)(23)). Finally, we created a set of Venn diagrams comparing genes identified using METRO with those identified in the TWAS that were conducted as part of the WMH(22) and AD (EA GWAS)(23) input GWAS studies. We used the *geneSynonym* R package(42) to ensure that genes named differently across studies were captured.

#### 2.3.4 Functional Enrichment Analysis

To characterize the biological function of the identified genes by METRO for general cognitive function, WMH and AD, we performed gene set enrichment analysis. Specifically, we used the g:GOSt(43) tool on the web software g:Profiler and mapped the genes to known functional informational sources, including Gene Ontology (GO): molecular function (MF), GO: biological process (BP), GO: cellular component (CC), Kyoto Encyclopedia of Genes and Genomes (KEGG), Reactome (REAC), WikiPathways (WP), Transfac (TF), MiRTarBase (MIRNA), Human Protein Atlas (HPA), CORUM protein complexes, and Human Phenotype Ontology (HP). In this analysis, we used the default option g:SCS method (Set Counts and Sizes) in g:Profiler for multiple testing correction and presented pathways identified with an adjusted p-value < 0.05. Driver terms in GO are highlighted using a two-stage algorithm for filtering GO enrichment results, providing a more efficient and reliable approach compared to traditional clustering methods. This feature groups significant terms into sub-ontologies based on their relations, and the second stage identifies leading gene sets that give rise to other significant functions in the same group of terms. This method uses a greedy search strategy that recalculates hypergeometric p-values and results in the consideration of multiple leading terms in a component, rather than selection of terms with the highest significance level.

## Results

In Table 1, we provide descriptive statistics for the samples used in the eQTL mapping study (e.g., 1,032 AA and 801 EA from GENOA) and the four input GWAS.(21–23) The GENOA eQTL study included participants with a mean age of 56.9 (SD=10.0) years. More than half of participants were female (65.6%). Mean age of participants was 56.9 (SD=7.8) years in the general cognitive function GWAS(21), and 64.2 years in the WMH GWAS.(22) In the AD GWAS in EA (23), mean age was 73.6 (SD=8.1) years for cases and 67.9 (SD=8.6) years for controls. In the AD GWAS in AA (24), mean age was 74.2 (SD=13.6) years for all participants.

Using METRO, we identified 602 genes associated with general cognitive function, 45 genes associated with WMH, 231 genes associated with AD (EA GWAS), and 9 genes associated with AD (AA GWAS) that were significant at the Bonferroni corrected alpha level (P<2.90−10^-6^; Figure 1, Tables S1-3). Genomic inflation factors for the TWAS p-values ranged from 1.00 to 2.55 (Figure 2). Among the three neurocognitive outcomes, prior to fine-mapping, METRO TWAS identified the *ICA1L* gene overlapping between WMH and AD (from EA GWAS); the *FMNL1* gene overlapping between WMH and general cognitive function; and 22 genes enriched in AD-related pathways and functions overlapping between general cognitive function and AD (from EA GWAs) (Figure 3a; Figure S1). After fine-mapping, the only overlapping gene that remained was *ICA1L* between WMH and AD (Figure 3b). The METRO TWAS for AD (AA GWAS) identified 9 genes overlapping with those identified in AD (EA GWAS); however, following fine-mapping, only *TOMM40* overlapped between the two AD TWASs (Figure 3b).

**Figure 1.**
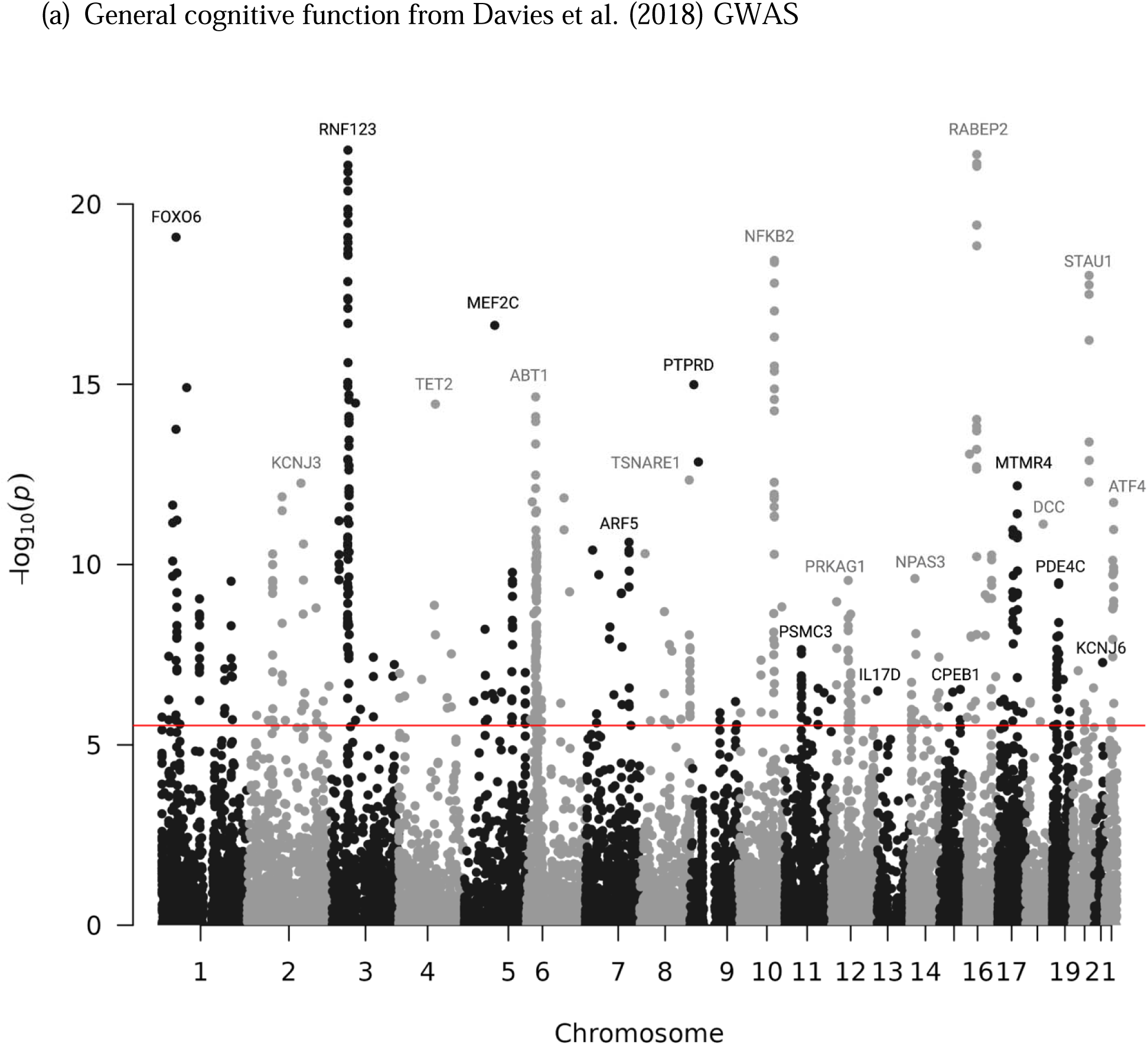
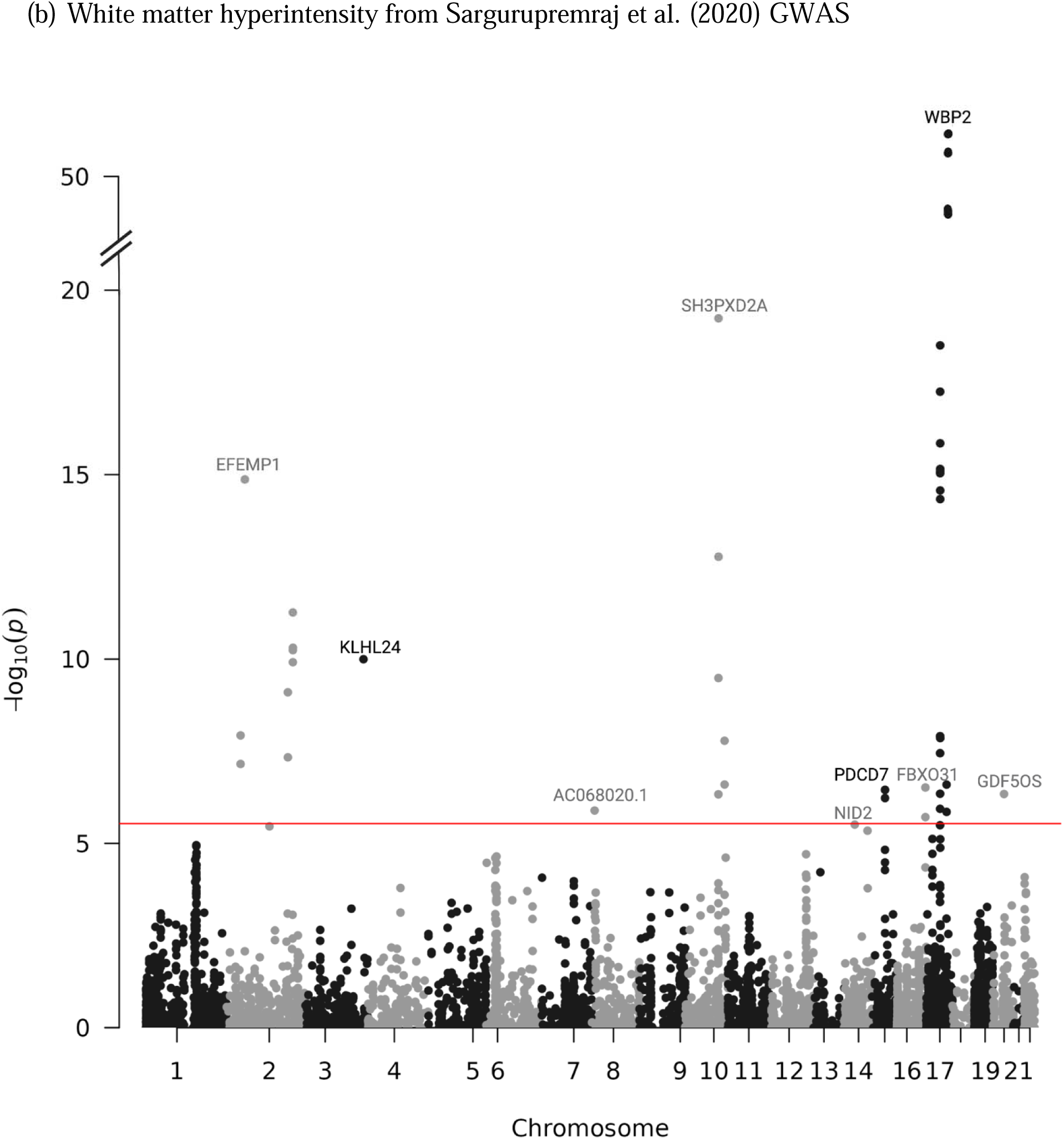
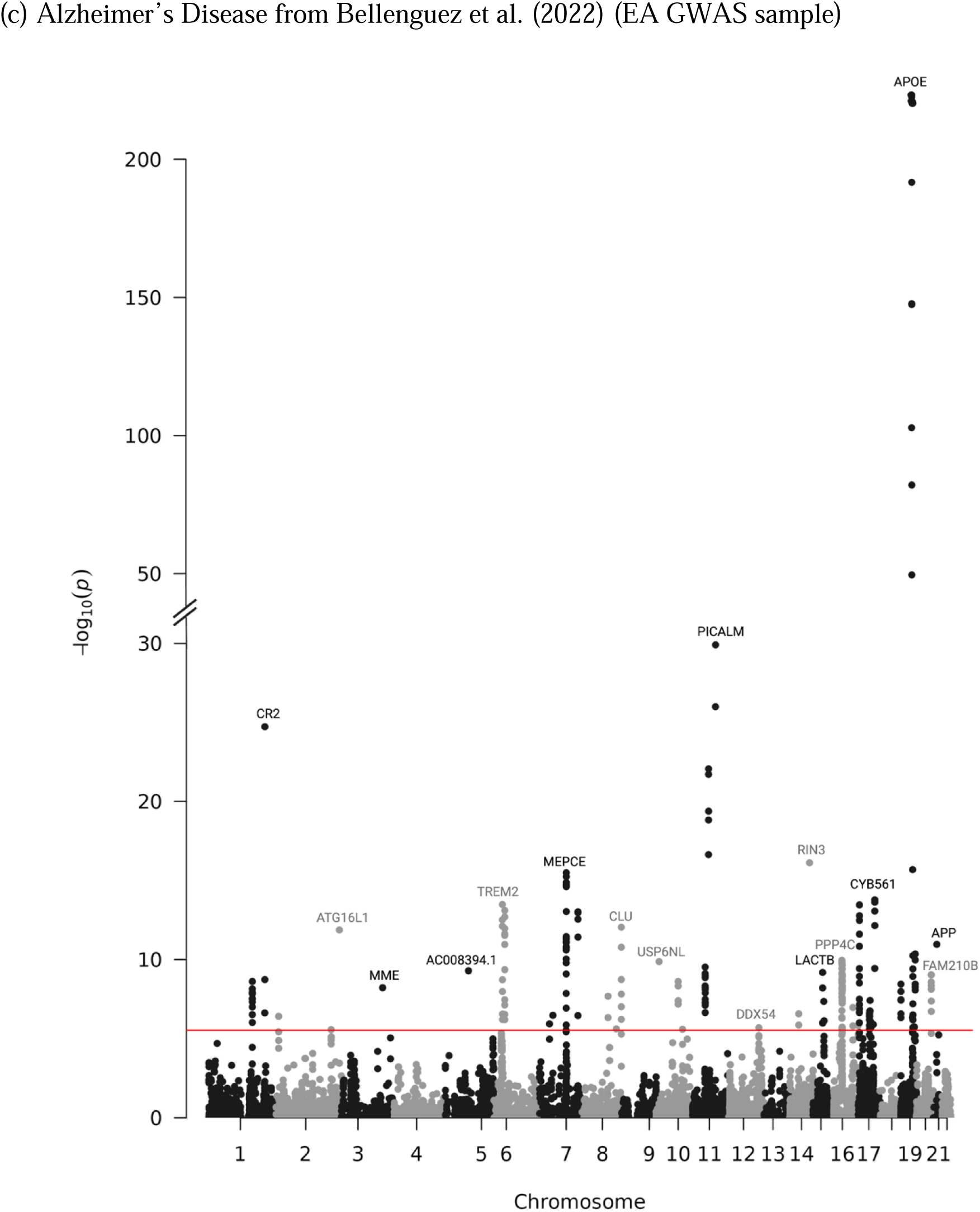
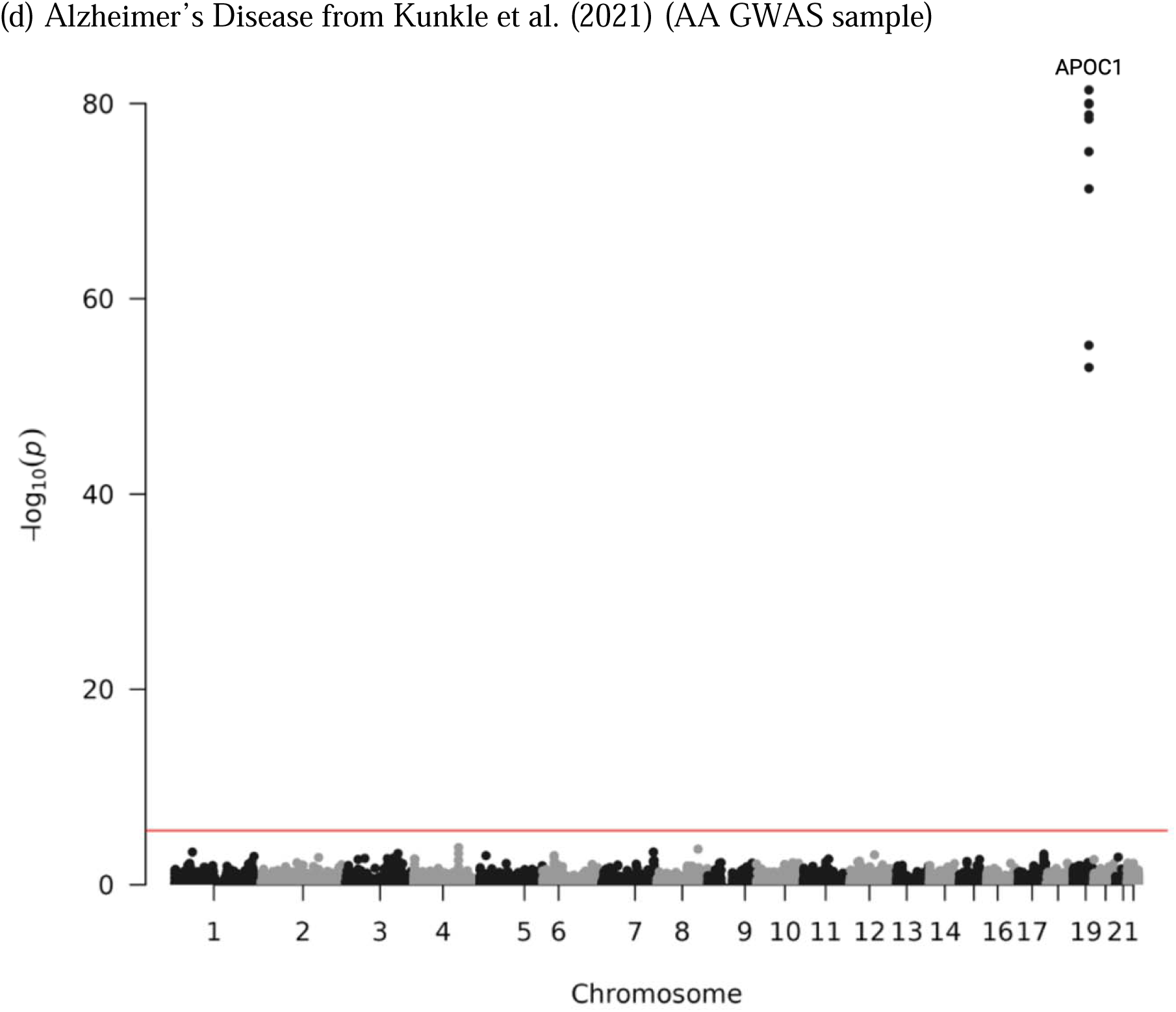
Manhattan plots of -log_10_ p-values for gene-trait associations in METRO. Manhattan plots of the association between genes and (a) general cognitive function using summary statistics from Davies et al. (2018), (b) White matter hyperintensity from Sargurupremraj et al. (2020), (c) Alzheimer’s disease from Bellenguez et al. (2022) (EA GWAS sample) and (d) Alzheimer’s disease from Kunkle et al. (2021) (AA GWAS sample) using GENOA gene expression data. The red line indicates significance after Bonferroni correction (P<2.90−10^-6^).

**Figure 2.**
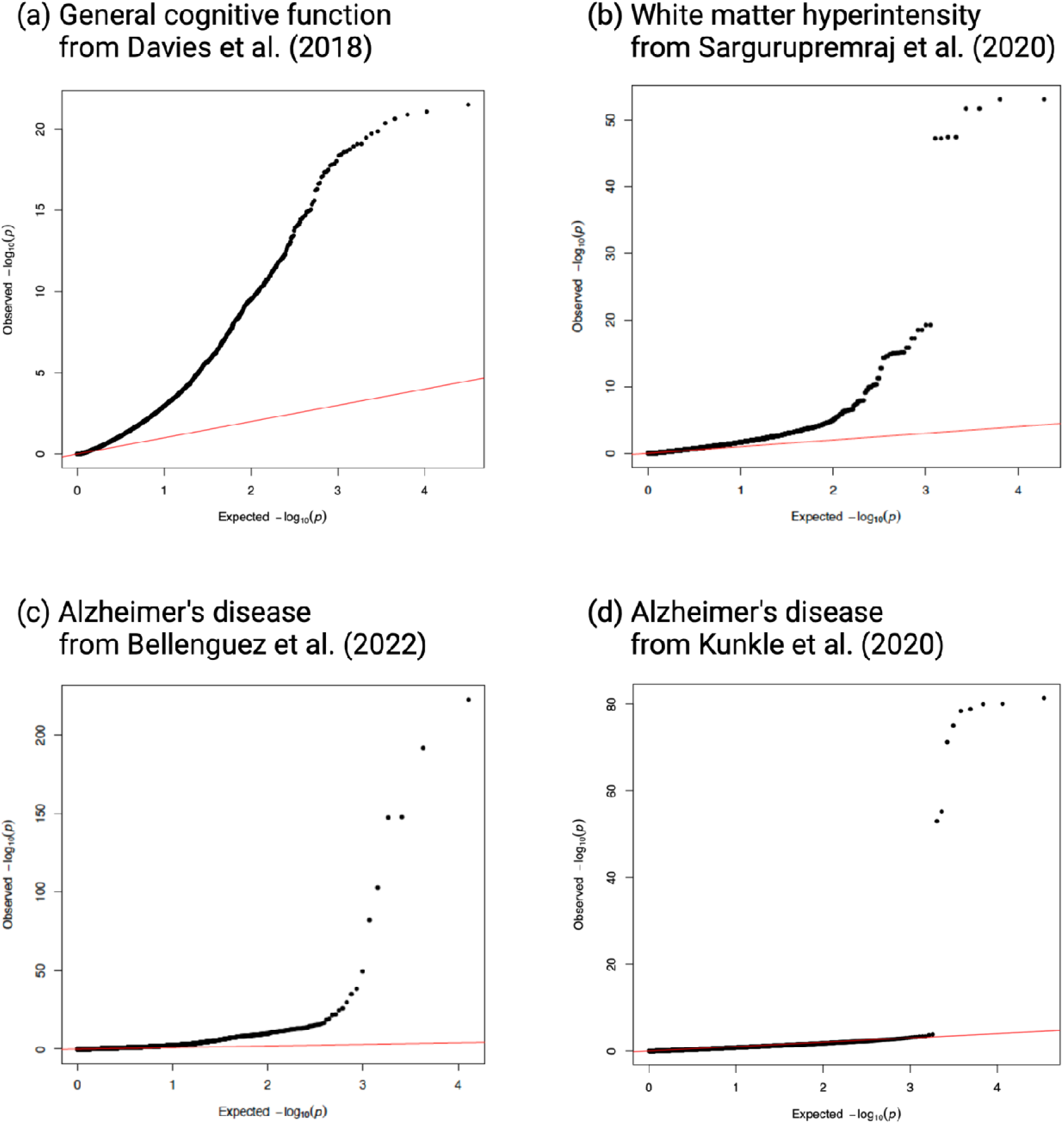
Quantile-quantile plots of -log_10_ p-values for gene-trait associations in METRO. Q-Q plots of the associations between genes and (a) general cognitive function (λ= 2.55) using summary statistics from Davies et al. (2018), (b) white matter hyperintensity (λ= 1.45) from Sargurupremraj et al. (2020), (c) Alzheimer’s disease (λ= 2.09) from Bellenguez et al. (2022) (EA GWAS sample) and (d) Alzheimer’s disease (λ= 1.0) from Kunkle et al. (2021)(AA GWAS sample) using GENOA gene expression data.

**Figure 3.**
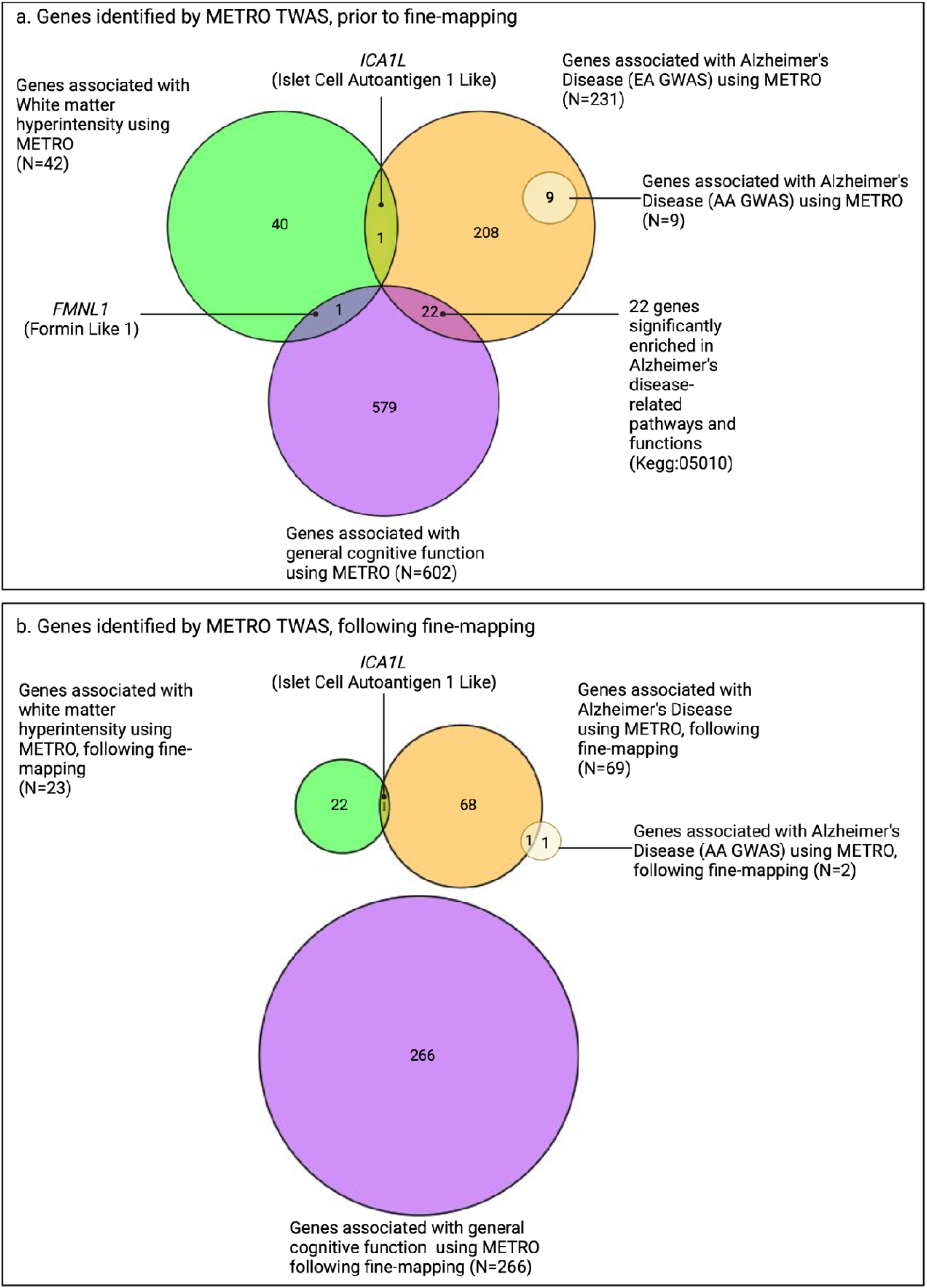
Venn diagrams comparing number of genes associated with general cognitive function, white matter hyperintensity and Alzheimer’s disease (AD) in European ancestry (EA) and AD in African ancestry (AA) using METRO, prior to and following FOCUS fine-mapping. Venn diagrams comparing the number of genes associated with general cognitive function (purple; N=266 genes), white matter hyperintensity (WMH; green; N=23 genes) Alzheimer’s disease (AD) in EA (orange; N=69 genes), and AD in AA (yellow; N=2 genes), (a) prior to fine-mapping and (b) following FOCUS(5) fine-mapping using METRO and GENOA expression data after Bonferroni correction (P<2.90−10^-6^), with GWAS summary statistics obtained from the Davies et al. (2018), Sargurupremraj et al. (2020), Bellenguez et al. (2022), and Kunkle et al. (2021).

For all identified genes, we also examined the contribution weights of expression prediction models for the EA and AA ancestries, prior to fine-mapping (P<2.90−10^-6^; Figure 4). For the WMH TWAS, we found that a large proportion of genes had stronger contributions from EA weights than AA weights (65.2%). This is consistent with Li et al. (2022)(20) who found that the gene expression prediction models constructed in the same ancestry as the input GWAS, in this case EA, often have larger contribution weights than those constructed in other ancestries. However, for both general cognitive function and AD (EA and AA GWAS), the contributions from EA and AA weights were similar, which likely increased power to identify genes relevant to AA.

**Figure 4.**
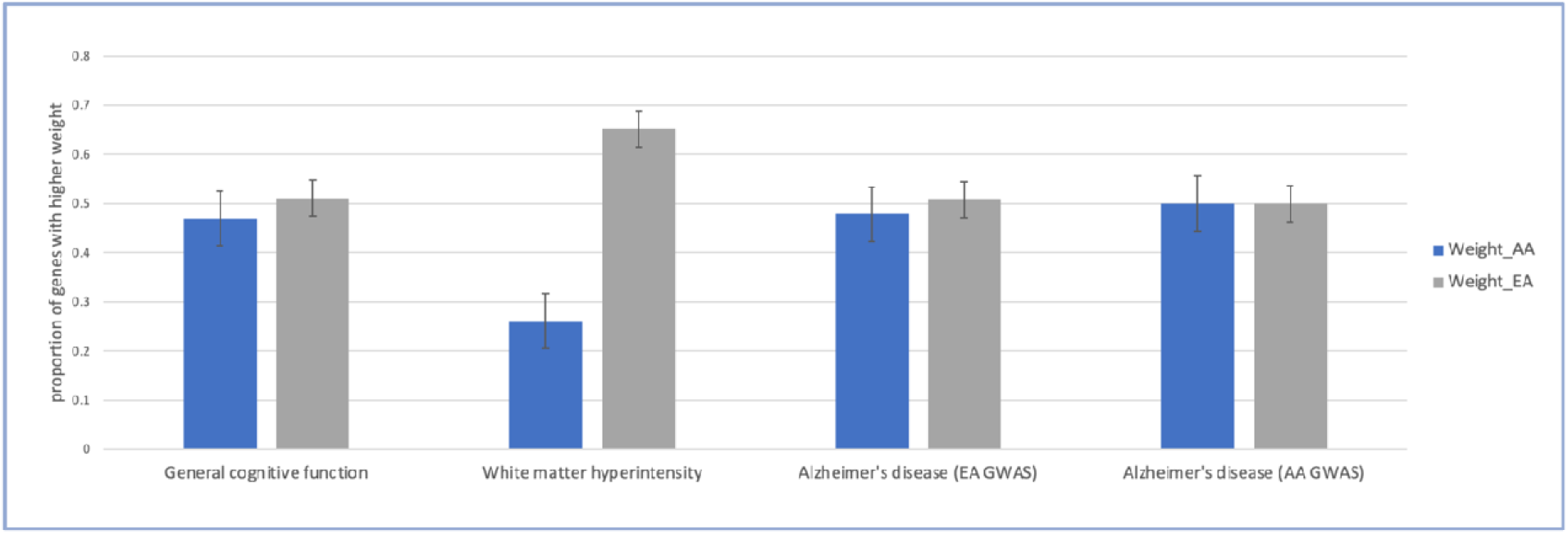
Contribution weights of expression prediction models across all significant fine-mapped genes identified by METRO. Barplots of general cognitive function, white matter hyperintensity and Alzheimer’s disease (AD) in European ancestry (EA) and AD in African ancestry (AA) comparing the proportion of significant genes with higher contribution weights of expression prediction models across all significant genes (P<2.90−10^-6^). Black bars are the standard errors for the estimated proportions.

After fine-mapping, there were 266 genes in the 90%-credible set across 172 different genomic regions for general cognitive function. This gene set included 82 genes that were not previously identified in the SNP-based GWAS results (mapped to the nearest gene) or the gene-based analysis results from Davies et al. (2018)(21) (Figure 5, Table S1); however, it is likely that some of these genes are in broader genomic regions tagged by the GWAS-identified SNPs. Specifically, there were 126 and 168 overlapping genes between METRO and the SNP-based and gene-based associations from Davies et al. (2018)(21), respectively (Figure 5). The 266 METRO-identified genes were enriched in regulatory pathways involved in protein binding (p_adj_ = 1.17 x 10^-5^), developmental cell growth (p_adj_ = 3.33 x 10^-5^), and protein metabolic process (p_adj_ = 7.18 x 10^-4^), as well as neurodevelopmental processes such as neuron to neuron synapse (p_adj_ = 1.22 x 10^-3^) and neuron projection (p_adj_ = 7.14 x 10^-3^; Figure 6). The 82 genes that were not previously identified in Davies et al. (2018)(21) were enriched for positive regulation of biological process (p_adj_ = 1.77 x 10^-2^), proteasome activator complex (p_adj_ = 1.00 x 10^-2^), nucleoplasm (p_adj_ = 1.29 x 10^-2^) and chromatin (p_adj_ = 4.71 x 10^-5^; Figure S2).

**Figure 5.**
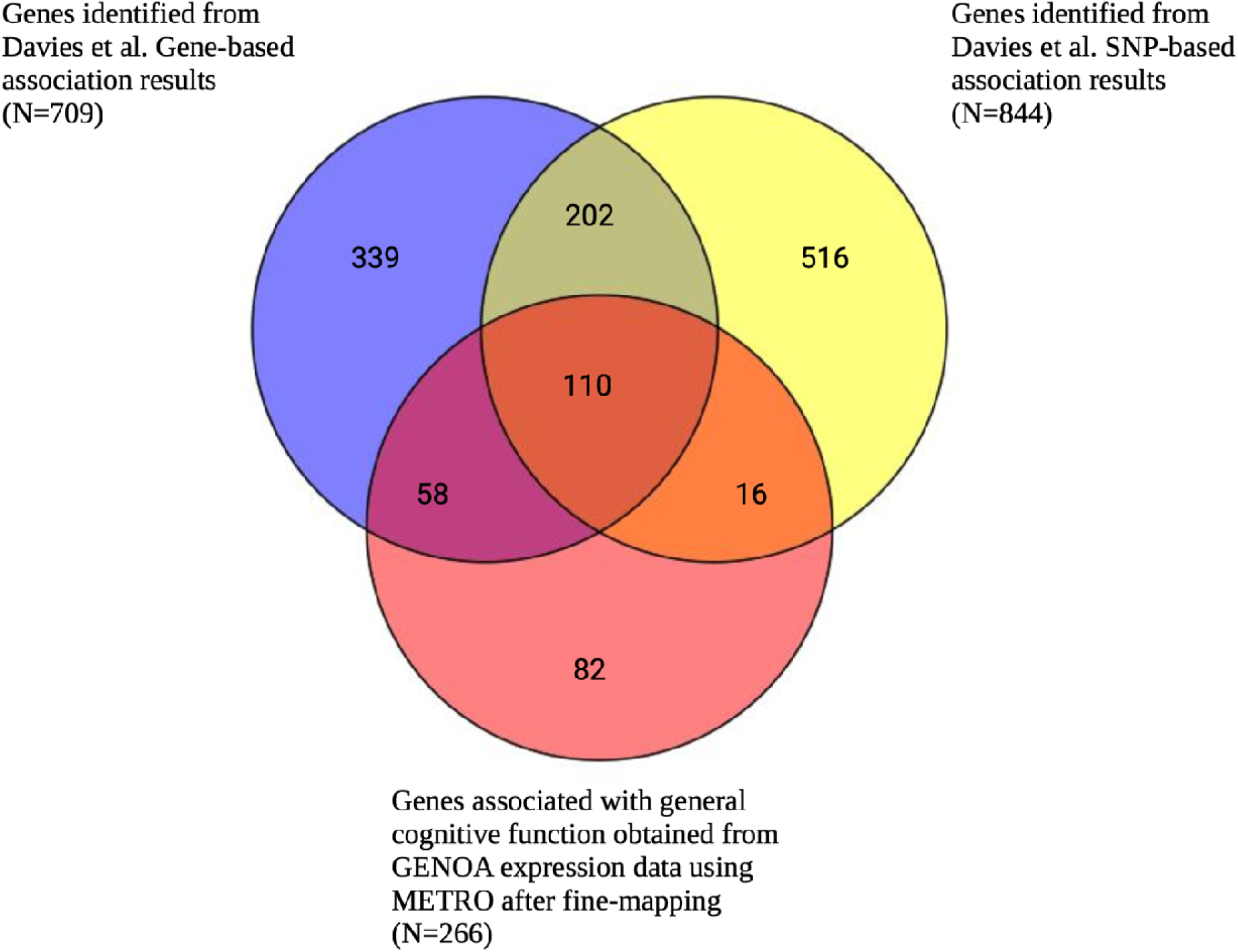
Venn diagram comparing number of METRO-identified genes associated with general cognitive function following FOCUS fine-mapping and genes identified by Davies et al. (2018) gene-based and SNP-based analyses. Venn diagram comparing the number of genes associated with general cognitive function obtained from METRO using GENOA gene expression data after Bonferroni correction (P<2.90−10^-6^) and FOCUS fine-mapping (red) and Davies et al. (2018). Davies et al. results included SNP-based association results that were mapped to the nearest gene (P<5−10^-8^; yellow), and gene-based association results (P<2.75−10^-6^; blue).

**Figure 6.**
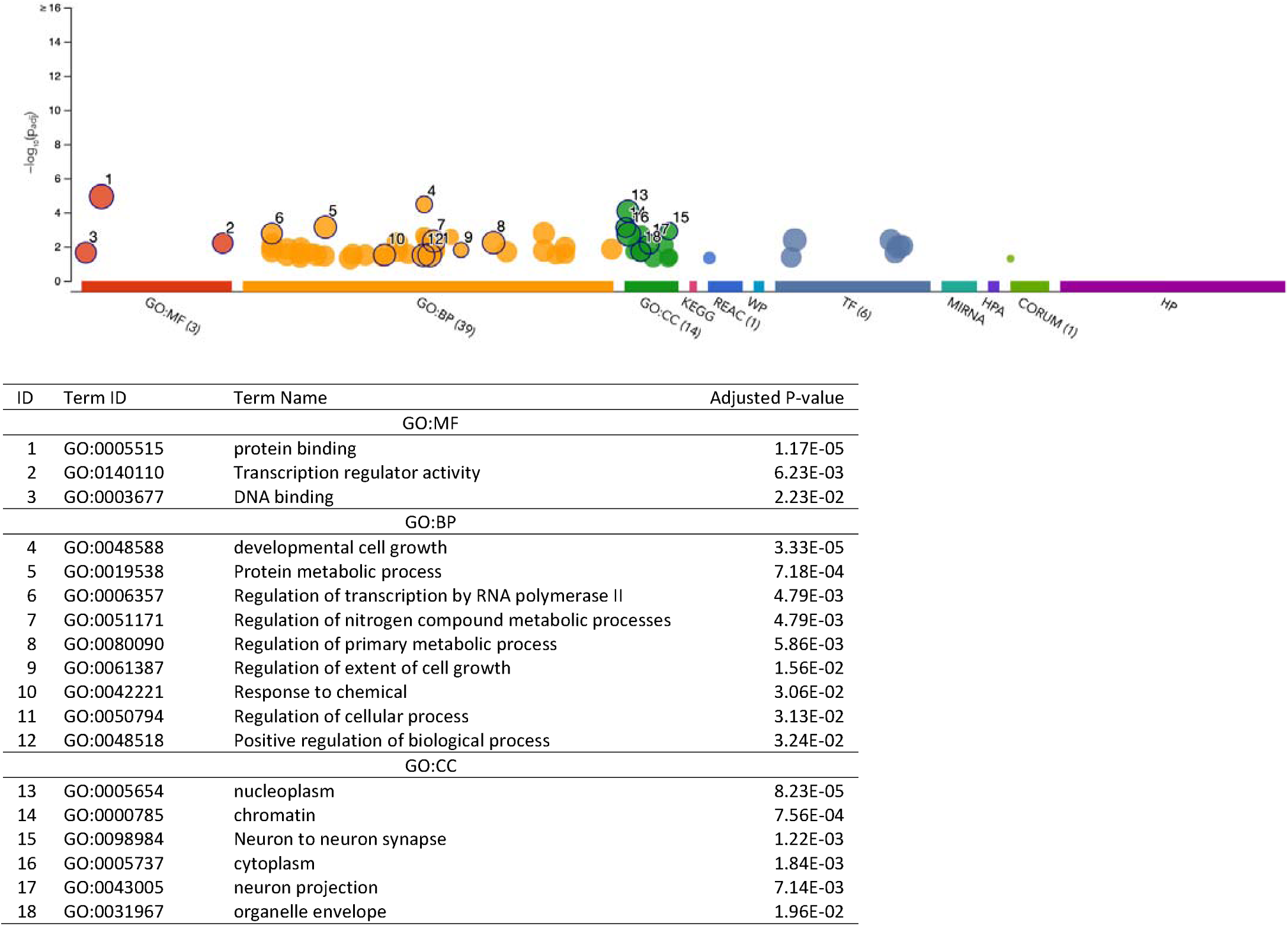
Functional enrichment analysis on the fine-mapped gene set identified for general cognitive function using METRO TWAS (N=266 genes). The top panel consists of a Manhattan plot that illustrates the enrichment analysis results. The x-axis represents functional terms that are grouped and color-coded by data sources, including Gene Ontology (GO): molecular function (MF; red), GO: biological process (BP; orange), GO: cellular component (CC; dark green), Kyoto Encyclopedia of Genes and Genomes (KEGG; pink), Reactome (REAC; dark blue), WikiPathways (WP; turquoise), Transfac (TF; light blue), MiRTarBase (MIRNA; emerald green), Human Protein Atlas (HPA; dark purple), CORUM protein complexes (light green), and Human Phenotype Ontology (HP; violet), in order from left to right. The y-axis shows the adjusted enriched -log_10_ p-values <0.05. Multiple testing correction was performed using g:SCS method (Set Counts and Sizes) that takes into account overlapping terms. The top panel highlights driver GO terms identified using the greedy filtering algorithm in g:Profiler. The light circles represent terms that were not significant after filtering. The circle sizes are in accordance with the corresponding term size (i.e., larger terms have larger circles). The number in parentheses following the source name in the x-axis shows how many significantly enriched terms were from this source.

After fine-mapping, there were 23 genes in the 90%-credible set across 15 genomic regions for WMH, including 12 genes that were not previously identified in the SNP-based GWAS results mapped to the nearest gene or the gene-based analysis results from Sargurupremraj et al. (2020)(22) (Figure 7, Table S2). Specifically, there were 7 and 12 overlapping genes between METRO and the SNP-based and gene-based associations from Sargurupremraj et al. (2020)(22), respectively (Figure 7). The 23 METRO-identified genes were enriched for zinc finger motif (p_adj_ = 1.27 x 10^-2^), miRNA has-212-5p (p_adj_ = 1.94 x 10^-2^) and retinal inner plexiform layer (p_adj_ = 3.86 x 10^-2^; Figure 8). The 12 genes associated with WMH that were previously not identified by Sargurupremraj et al. (2020)(22) were enriched for DNA binding domain Zinc Finger Protein 690 (ZNF690; p_adj_ = 2.52 x 10^-3^) and ClpX protein degradation complex (p_adj_ = 4.97 x 10^-2^; Figure S3).

**Figure 7.**
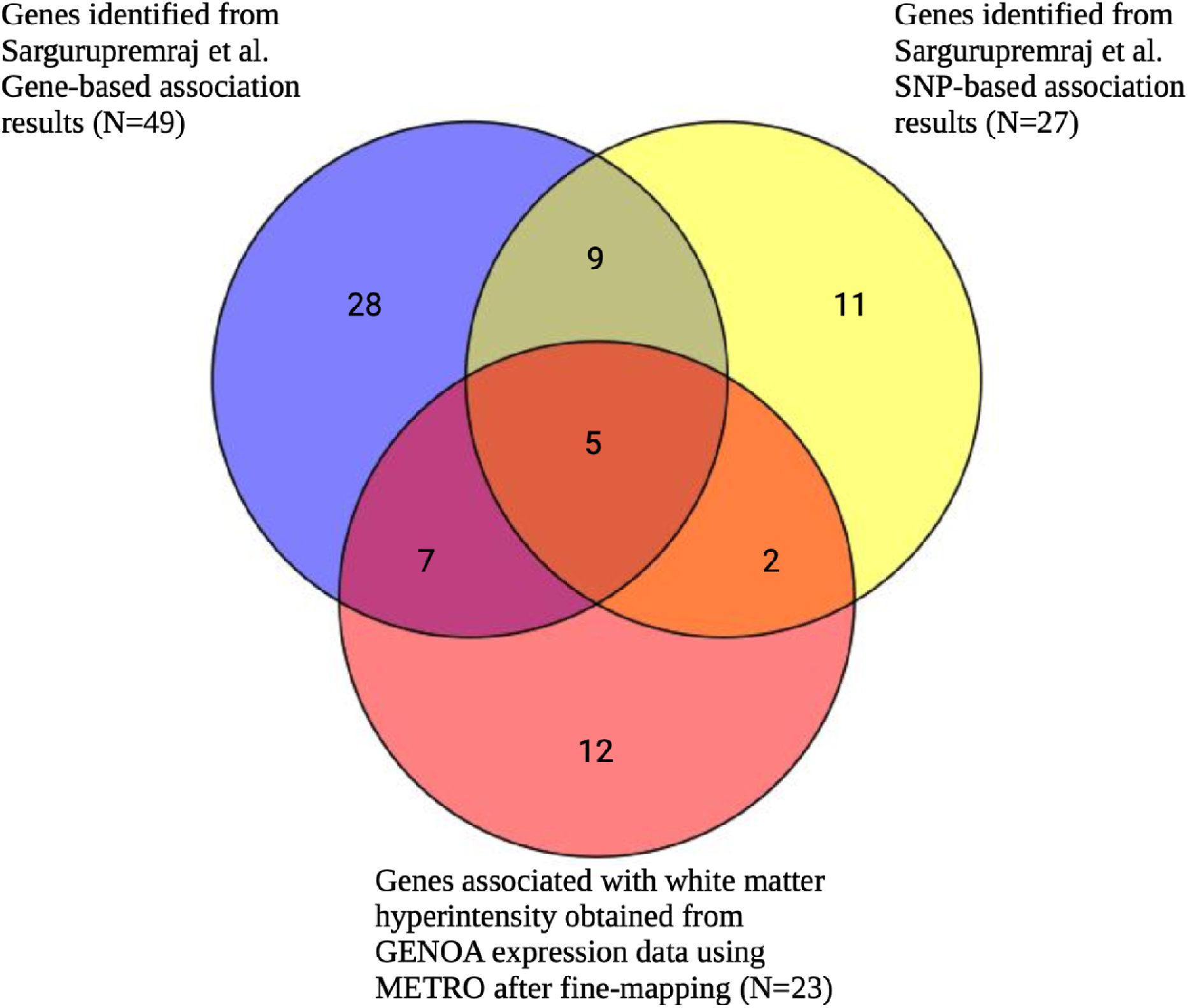
Venn diagram comparing number of METRO-identified genes associated with white matter hyperintensity following FOCUS fine-mapping and genes identified by Sargurupremraj et al. (2020) gene-based and SNP-based analyses. Venn diagram comparing the number of significantly associated genes associated with white matter hyperintensity (WMH) obtained from METRO using GENOA expression data after Bonferroni correction (P<2.90−10^-6^), and fine-mapping (red) and Sargurupremraj et al. (2020). Sargurupremraj et al. results included SNP-based association results that were mapped to the nearest gene (P<5−10^-8^; yellow), and gene-based association results (P<2.77−10^-6^; blue).

**Figure 8.**
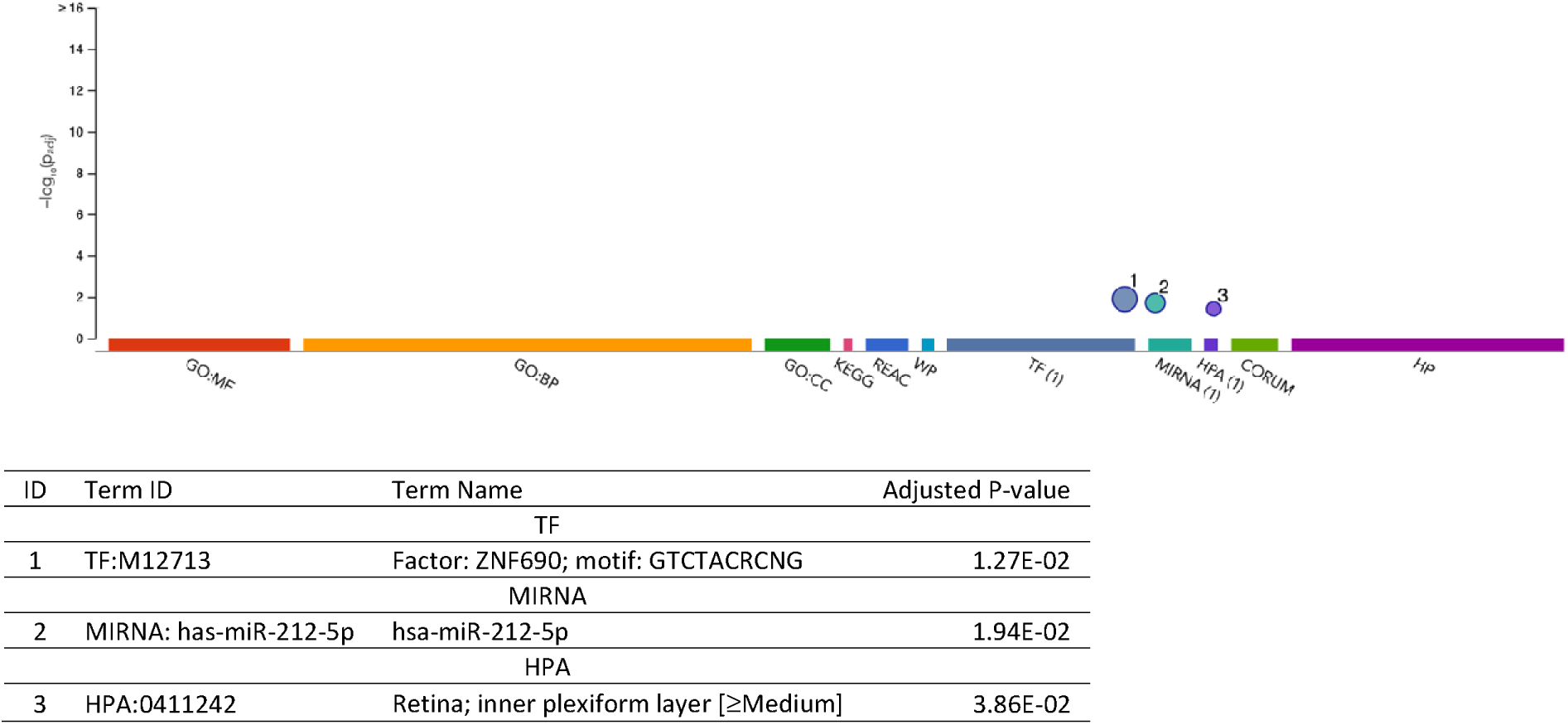
Functional enrichment analysis on the fine-mapped gene set identified for white matter hyperintensity using METRO TWAS (N=23 genes). The top panel consists of a Manhattan plot that illustrates the enrichment analysis results. The x-axis represents functional terms that are grouped and color-coded by data sources, including Gene Ontology (GO): molecular function (MF; red), GO: biological process (BP; orange), GO: cellular component (CC; dark green), Kyoto Encyclopedia of Genes and Genomes (KEGG; pink), Reactome (REAC; dark blue), WikiPathways (WP; turquoise), Transfac (TF; light blue), MiRTarBase (MIRNA; emerald green), Human Protein Atlas (HPA; dark purple), CORUM protein complexes (light green), and Human Phenotype Ontology (HP; violet), in order from left to right. The y-axis shows the adjusted enriched -log_10_ p-values < 0.05. Multiple testing correction was performed using g:SCS method (Set Counts and Sizes) that takes into account overlapping terms. The top panel highlights driver GO terms identified using the greedy filtering algorithm in g:Profiler. The light circles represent terms that were not significant after filtering. The circle sizes are in accordance with the corresponding term size (i.e., larger terms have larger circles). The number in parentheses following the source name in the x-axis shows how many significantly enriched terms were from this source.

After fine-mapping, there were 69 genes in the 90%-credible set across 56 genomic regions associated with AD (EA GWAS), including 45 genes that were not previously identified in the SNP-based GWAS results mapped to the nearest gene or the gene prioritization analysis results from Bellenguez et al. (2022)(23) (Figure 9, Table S3). Specifically, there were 16 and 14 overlapping genes between METRO and the SNP-based and gene prioritization test results from Bellenguez et al. (2022)(23), respectively (Figure 9). The 69 METRO-identified genes were enriched for AD-associated processes including regulation of amyloid fibril formation (p_adj_ = 1.87 x 10^-3^), amyloid-beta clearance (p_adj_ = 1.90 x 10^-3^), microglial cell activation (p_adj_ = 5.79 x 10^-3^), amyloid-beta metabolic process (p_adj_ = 1.07 x 10^-2^), and neurofibrillary tangle (p_adj_ = 2.80 x 10^-4^; Figure 10). The 45 genes associated with AD that were previously not identified by Bellenguez et al. (2022)(23) were enriched for hematopoietic cell lineage (p_adj_ = 1.73 x 10^-3^) and neurofibrillary tangle (p_adj_ = 9.13 x 10^-3^; Figure S4).

**Figure 9.**
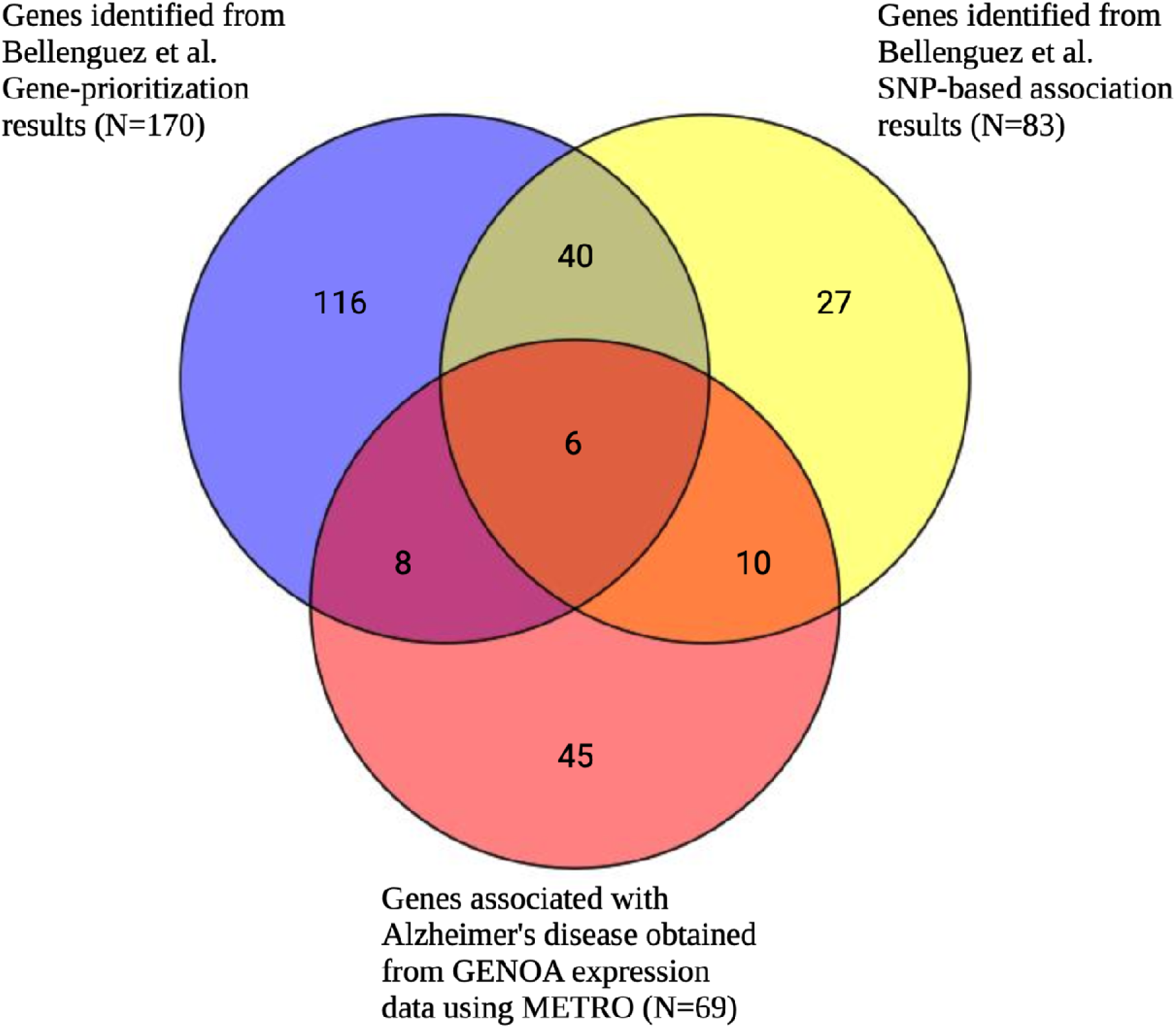
Venn diagram comparing number of METRO-identified genes associated with Alzheimer’s disease (EA GWAS) following FOCUS fine-mapping and genes identified by Bellenguez et al. (2022) gene prioritization and SNP-based analyses. Venn diagram comparing the number of significantly associated genes associated with Alzheimer’s disease (from EA GWAS) obtained from METRO using GENOA expression data after Bonferroni correction (P<2.90−10^-6^) and fine-mapping (red) and Bellenguez et al. (2022). Bellenguez et al. results included SNP-based association results that were mapped to the nearest gene (P<5−10^-8^; yellow), and gene prioritization results for the genes in the novel AD risk loci (blue). In the gene prioritization analysis, Bellenguez et al. analyzed the downstream effects of new AD-associated loci on molecular phenotypes (i.e., expression, splicing, protein expression, methylation and histone acetylation) in various *cis*-quantitative trait loci (*cis*-QTL) catalogues from AD-relevant tissues, cell types and brain regions.

**Figure 10.**
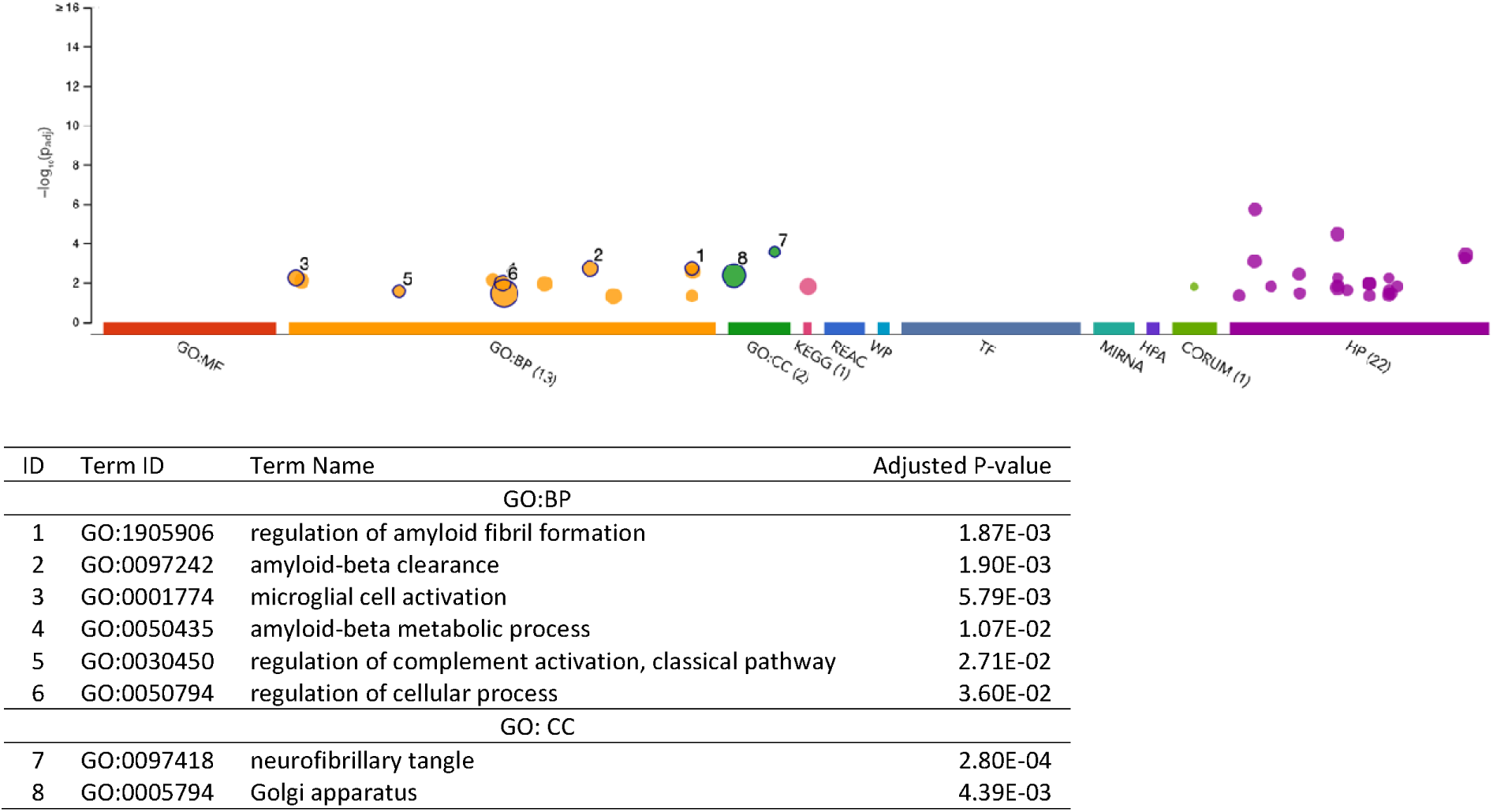
Functional enrichment analysis on the fine-mapped gene set identified for Alzheimer’s disease (EA GWAS) using METRO TWAS (N=69 genes). The top panel consists of a Manhattan plot that illustrates the enrichment analysis results. The x-axis represents functional terms that are grouped and color-coded by data sources, including Gene Ontology (GO): molecular function (MF; red), GO: biological process (BP; orange), GO: cellular component (CC; dark green), Kyoto Encyclopedia of Genes and Genomes (KEGG; pink), Reactome (REAC; dark blue), WikiPathways (WP; turquoise), Transfac (TF; light blue), MiRTarBase (MIRNA; emerald green), Human Protein Atlas (HPA; dark purple), CORUM protein complexes (light green), and Human Phenotype Ontology (HP; violet), in order from left to right. The y-axis shows the adjusted enriched -log_10_ p-values < 0.05. Multiple testing correction was performed using g:SCS method (Set Counts and Sizes) that takes into account overlapping terms. The top panel highlights driver GO terms identified using the greedy filtering algorithm in g:Profiler. The light circles represent terms that were not significant after filtering. The circle sizes are in accordance with the corresponding term size (i.e., larger terms have larger circles). The number in parentheses following the source name in the x-axis shows how many significantly enriched terms were from this source.

We identified 2 genes, *APOE* and *PVRL2*, in the 90%-credible set associated with AD (AA GWAS) (Table S4). After fine-mapping, none of these genes overlapped with SNP-based GWAS results mapped to the nearest gene or the gene-based analysis results from Kunkle et al. (2021), since they are both in the broader *APOE* region, which was the only identified gene in Kunkle et al (2021). The 2 METRO-identified genes were enriched for coreceptor-mediated virion attachment to host (p_adj_ = 4.96 x 10^-2^; Figure 11).

**Figure 11.**
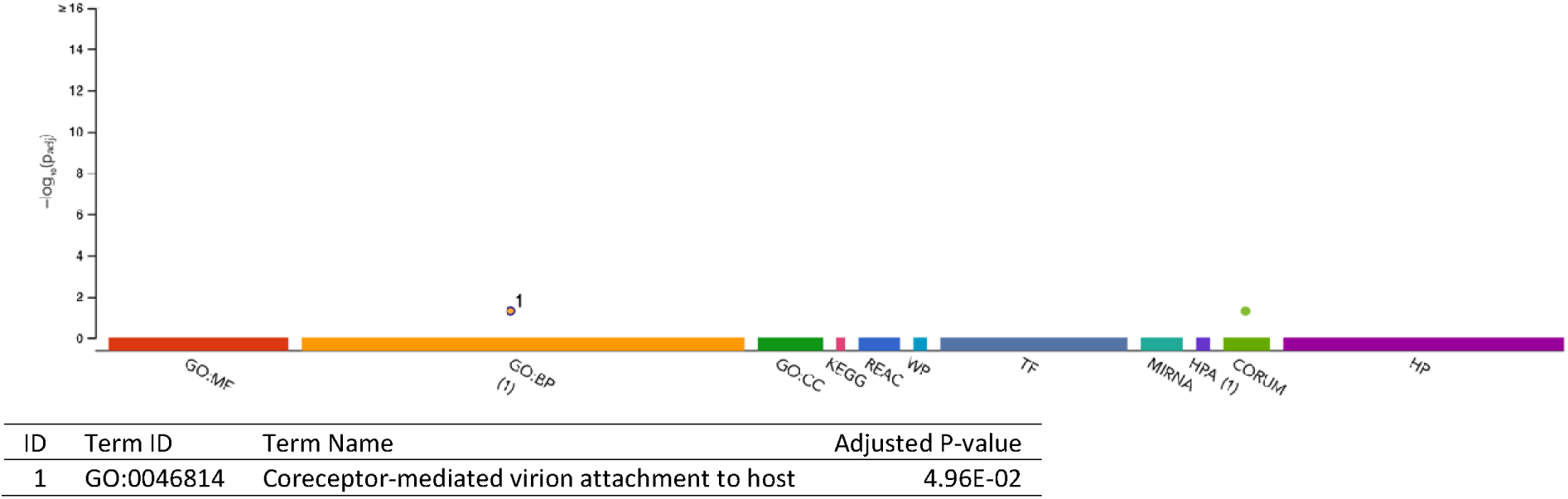
Functional enrichment analysis fine-mapped gene set identified for Alzheimer’s disease (AA GWAS) using METRO TWAS (N=2 genes). The top panel consists of a Manhattan plot that illustrates the enrichment analysis results. The x-axis represents functional terms that are grouped and color-coded by data sources, including Gene Ontology (GO): molecular function (MF; red), GO: biological process (BP; orange), GO: cellular component (CC; dark green), Kyoto Encyclopedia of Genes and Genomes (KEGG; pink), Reactome (REAC; dark blue), WikiPathways (WP; turquoise), Transfac (TF; light blue), MiRTarBase (MIRNA; emerald green), Human Protein Atlas (HPA; dark purple), CORUM protein complexes (light green), and Human Phenotype Ontology (HP; violet), in order from left to right. The y-axis shows the adjusted enriched -log_10_ p-values < 0.05. Multiple testing correction was performed using g:SCS method (Set Counts and Sizes) that takes into account overlapping terms. The top panel highlights driver GO terms identified using the greedy filtering algorithm in g:Profiler. The light circles represent terms that were not significant after filtering. The circle sizes are in accordance with the corresponding term size (i.e., larger terms have larger circles). The number in parentheses following the source name in the x-axis shows how many significantly enriched terms were from this source.

We compared the genes identified by METRO before and after fine-mapping with those identified by TWAS studies in Sargurupremraj et al. (2020)(22) and Bellenguez et al. (2022)(23) which used TWAS-Fusion (Figure 12). For WMH, there were 16 and 10 genes identified both by METRO before and after fine-mapping and by the TWAS-Fusion analysis conducted by Sargurupremraj et al. (2020)(22), respectively (Table 2). For AD, there were 24 and 10 genes identified both by METRO before and after fine-mapping and by the TWAS-Fusion followed by FOCUS fine-mapping analysis conducted by Bellenguez et al. (2022)(23) (Table 3). *ICA1L* was the only gene overlapping between all four AD and WMH TWAS association results.

**Figure 12.**
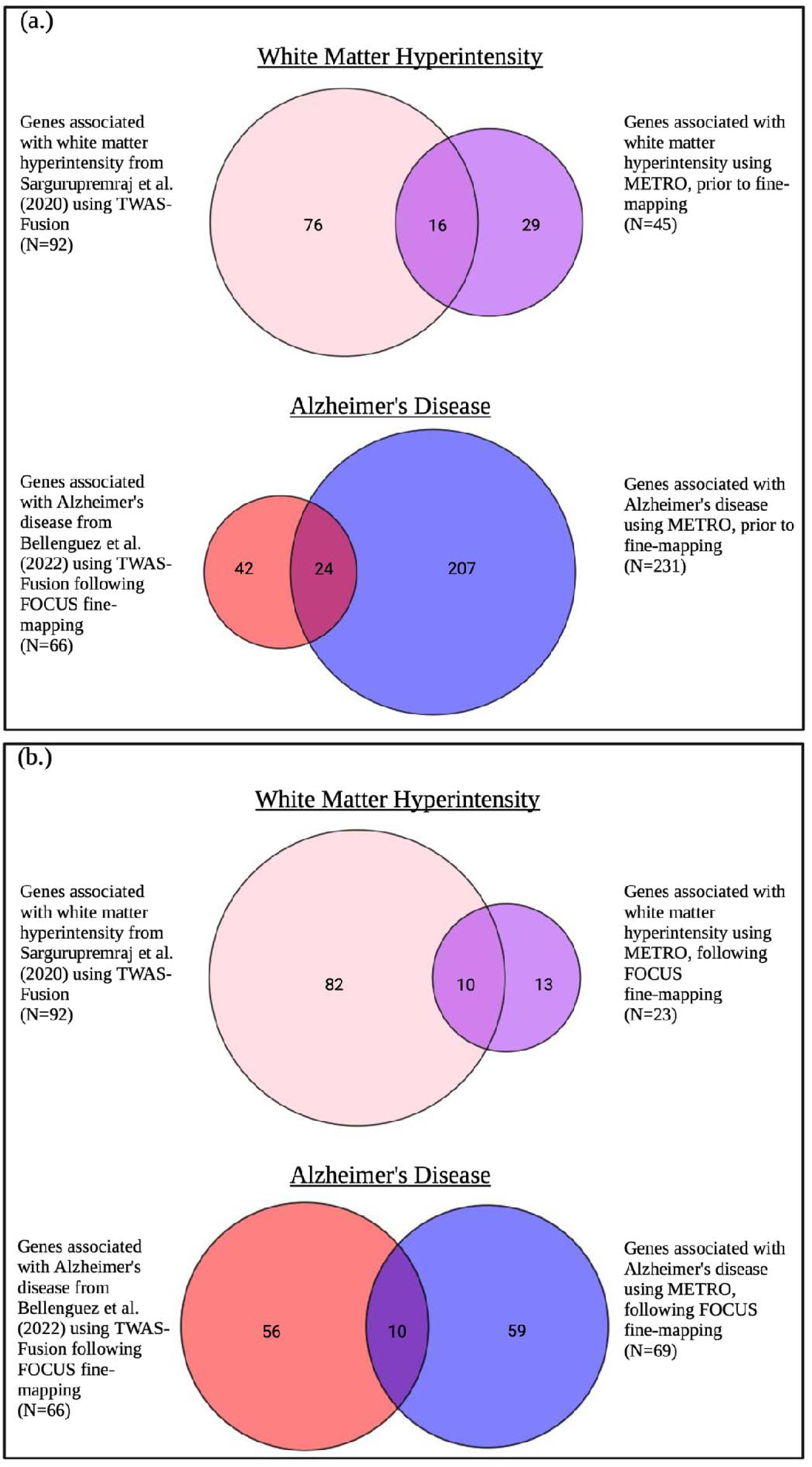
Venn diagram comparing METRO TWAS results prior to and following FOCUS fine-mapping with TWAS results from Sargurupremraj et al. (2020) and Bellenguez et al. (2022). Venn diagram comparing METRO TWAS results (a) prior to and (b) following FOCUS fine-mapping with TWAS results using Fusion for white matter hyperintensity from Sargurupremraj et al. (2020) without fine-mapping and Alzheimer’s disease from Bellenguez et al. (2022) (EA GWAS) with FOCUS fine-mapping.

**Table 2.**
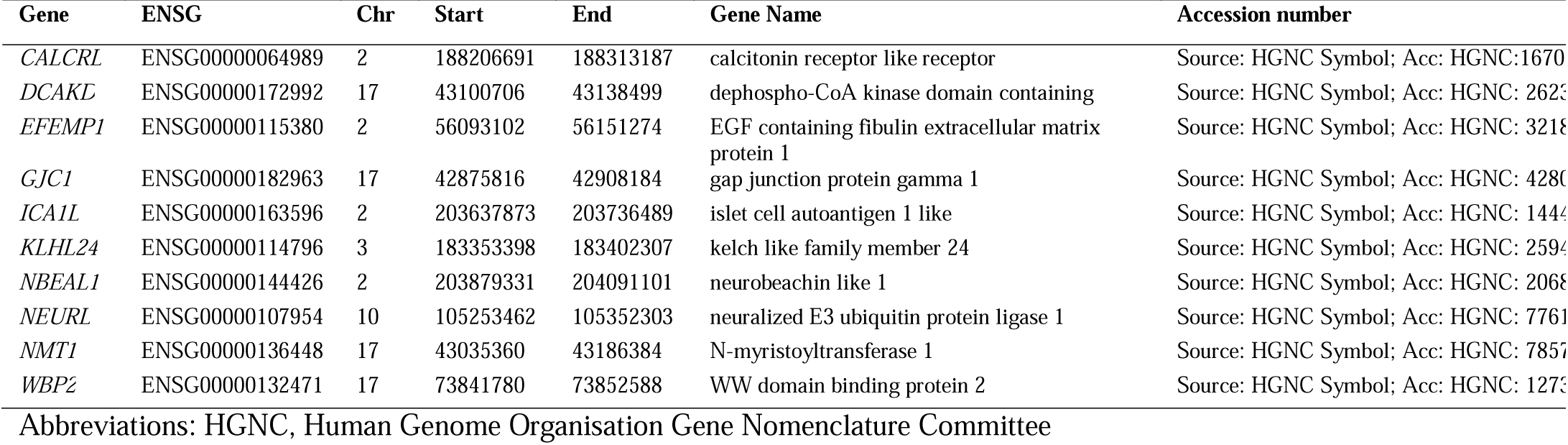
Genes for WMH identified both by METRO followed by fine-mapping with FOCUS and by TWAS-Fusion conducted by Sargurupremraj et al. (2020)

**Table 3.** Genes for AD identified both by METRO followed by fine-mapping with FOCUS and by TWAS-Fusion followed by fine-mapping with FOCUS conducted by Bellenguez et al. (2022)

| Gene | ENSG | Chr | Start | End | Gene Name | Accession number |
| --- | --- | --- | --- | --- | --- | --- |
| *BLNK* | ENSG00000095585 | 10 | 97948927 | 98031344 | B cell linker | Source:HGNC<br>Symbol;Acc:HGNC:14211 |
| *CPSF3* | ENSG00000119203 | 2 | 9563780 | 9613230 | cleavage and polyadenylation specific factor 3 | Source:HGNC<br>Symbol;Acc:HGNC:2326 |
| *DDX54* | ENSG00000123064 | 12 | 113594978 | 113623284 | DEAD-box helicase 54 | Source:HGNC<br>Symbol;Acc:HGNC:20084 |
| *GRN* | ENSG00000030582 | 17 | 42422614 | 42430474 | granulin precursor | Source:HGNC<br>Symbol;Acc:HGNC:4601 |
| *ICA1L* | ENSG00000163596 | 2 | 203637873 | 203736489 | islet cell autoantigen 1 like | Source:HGNC<br>Symbol;Acc:HGNC:14442 |
| *KLF16* | ENSG00000129911 | 19 | 1852398 | 1863578 | KLF transcription factor 16 | Source:HGNC<br>Symbol;Acc:HGNC:16857 |
| *LACTB* | ENSG00000103642 | 15 | 63414032 | 63434260 | lactamase beta | Source:HGNC<br>Symbol;Acc:HGNC:16468 |
| *PPP4C* | ENSG00000149923 | 16 | 30087299 | 30096697 | protein phosphatase 4 catalytic subunit | Source:HGNC<br>Symbol;Acc:HGNC:9319 |
| *SHARPIN* | ENSG00000179526 | 8 | 145153536 | 145163027 | SHANK associated RH domain interactor | Source:HGNC<br>Symbol;Acc:HGNC:25321 |
| *TBX6* | ENSG00000149922 | 16 | 30097114 | 30103245 | T-box transcription factor 6 | Source:HGNC<br>Symbol;Acc:HGNC:11605 |

Abbreviations: HGNC, Human Genome Organisation Gene Nomenclature Committee

## Discussion

While previous studies have identified genes associated with cognitive function, WMH, and AD, there are few TWAS that utilize genetic and gene expression data from multiple ancestries to elucidate gene-trait associations and molecular mechanisms underlying the etiologies of cognitive function and neurocognitive disorders. Using the METRO method in GWAS consisting primarily of EA followed by FOCUS fine-mapping, we identified 266, 23, and 69 genes associated with general cognitive function, WMH, and AD, respectively, with 82, 12 and 45 of them not previously identified in the original GWAS. In addition, using an AA GWAS, we identified 2 fine-mapped genes associated with AD, both of which are proximal to *APOE*. Studying the transcriptomic mechanisms underlying cognitive function, WMH and dementia using both EA and AA expression data may enhance our understanding of cognitive health prior to and following the onset of dementia and further allow us to generalize findings from large scale EA GWAS to other ancestries.

AD and SVD have overlapping features that contribute to dementia neuropathology including breakdown of the blood-brain barrier(44) and the presence of small cortical and subcortical infarcts, microbleeds, perivascular spacing, and WMH in brain tissue(45). After fine-mapping, Islet Cell Autoantigen 1 Like (*ICA1L*) was identified in both the WMH and AD TWAS. This is as a highly plausible prioritized gene that is likely to modulate the metabolism of amyloid precursor protein (APP)(23) and increase risk of AD. *ICA1L* encodes a protein whose expression is activated by type IV collagen and plays a crucial role in myelination(46). Increased *ICA1L* expression is also associated with lower risk of AD(47–49) and small vessel strokes (SVS), the acute outcomes of cerebral SVD, which may lead to VaD(50). Consistent with these studies, our TWAS found that decreased expression of *ICA1L* is associated with increased risk of AD and WMH, a subclinical indicator of SVD. Single-cell RNA-sequencing has shown *ICA1L* expression to be enriched in cortical glutamatergic excitatory neurons, which are crucial components in neural development and neuropathology through their role in cell proliferation, differentiation, survival, neural network formation and cell death(51,52). *ICA1L* has been examined as a possible drug target for SVD, AD, and other neurodegenerative diseases(50,53); however, it is not recommended as a prioritized drug at this time due to potential side effects including increased risk of coronary artery disease and myocardial infarction as well as lower diastolic blood pressure(53). Nevertheless, *ICA1L* may contribute to overlapping AD and VaD neuropathology, and it could be a potential target for therapeutics and/or preventative treatments for AD and VaD in the future if adverse events can be reduced.

Our TWAS of AD (from EA GWAS) identified 45 genes that were not identified in the SNP-based GWAS results mapped to the nearest gene or the gene-based analysis reported in Bellenguez et al. (2022)(23). The 45 genes were enriched for hematopoietic cell lineage, which are progenitors of red and white blood cells including those related to immunity (e.g., natural killer cells, T- and B-lymphocytes and other types of leukocytes)(54–60). Our TWAS identified genes that have been previously associated with AD, including *APOE, TOMM40, APOC4, CLU, PICALM* and *CR2,* among others(23,61,62). While we identified *APOE,* the strongest genetic risk factor for AD in most populations, after fine-mapping, we did not identify *ABCA7* which confers an equal or even greater risk for AD in AA(63–65). This finding is perhaps not surprising considering that our TWAS was conducted using an EA GWAS, and the strength of association between *ABCA7* and AD is comparatively weaker in EA than in AA.(65) To identify genes associated with AD risk in AA populations, specifically, it would be beneficial to perform a TWAS utilizing a well-powered AD GWAS in AA.

Our TWAS of AD (from AA GWAS) (24) identified only *TOMM40* and *PVRL2*, both proximal to *APOE*. *TOMM40* was also identified in our TWAS of AD (from EA GWAS), as well as other AD GWAS (23,61,62). *PVRL2* has been associated with metabolic syndrome, diabetic dyslipidemia, and AD(66,67). One study found that polymorphisms in *PVRL2* interact with variants in *TOMM40* to increase AD risk through pathways related to amyloid-beta metabolism in older Chinese adults(67). As larger AD GWAS in AA become available, we may be able to identify additional genes associated with disease while leveraging transcriptomic data from EA and AA.

In our AD EA TWAS, we also identified genes associated with other neurological and autoimmune diseases including Parkinson’s disease (*CYB561*(68) and *SLC25A39*(69)), Crohn’s disease (*ATG16L1*(70)), Amyotrophic lateral sclerosis (*SIGLEC9*(71)), and Riboflavin Transport Deficiency (*SLC52A1*(72)). These diseases have in common the progressive peripheral and cranial degeneration of neurons that impact processes such as voluntary muscle movement, vision, hearing and sensation. Although not explicitly identified in Bellenguez et al. (2022)(23), we also identified genes that were associated with AD in other studies including *RIN3* that is implicated in tau-mediated pathology, the *MS4A* (*4A* and *6A*) locus associated with mast cell activation, *TP53INP1* and *ZYX* that have been linked to myeloid enhancer activity(73), and *APOC4*, which is located proximal to *APOE*(74). We also identified additional genes involved in B cell autoimmunity (*HLA-DQA2*(75,76) and *CSTF1*(77)), neurodegenerative processes (*SUPT4H1*(78)*, C6orf10*(79)*, IKZF1*(80), and *DEDD*(81)), and neuronal growth (*IKZF1*(80) and *STYX*(82)). Our findings support the hypothesis that chronic activation of immune cells resident in the brain and peripheral nervous system appear to play a critical role in neuroinflammatory responses that drive the progression of neurodegeneration in AD.(83) Further, consistent with findings that AD and VaD often co-exist, our AD TWAS identified genes that were associated with lacunar and ischemic strokes as well as cerebral small vessel disease in other studies, including *SLC39A13*(84)*, RAPSN*(84)*, MAF1*(85), and *MME*(86,87).

Although our WMH TWAS identified 12 genes that were not included in the SNP-based GWAS results mapped to the nearest gene or the gene-based analysis reported in Sargurupremraj et al. (2020)(22), other studies found associations between *MAP1LC3B*(88)*, ARMS2*(89,90) and *HTRA1*(84) with ischemic stroke, lacunar stroke, and cerebral SVD. The WMH TWAS also identified genes associated with AD (*ARMS2*)(91), atrial fibrillation (*NEURL*(92) and *GJC1*(93)), innate immunity (*EFTUD2*(94)) and apoptosis and neurodevelopment (*PDCD7*(95)*, FBXO31*(96), and *ClpX*(97)). The 12 unique genes identified for WMH were enriched for DNA binding domain Zinc Finger Protein 690(98), which plays an essential role in gene regulation, transcription and various cellular processes, and ClpX protein degradation complex(99), which maintains protein homeostasis. Our findings were consistent with studies that showed neuroinflammation to be an immunological cascade reaction by glial cells of the central nervous system where innate immunity resides.

While our TWAS for general cognitive function did not show overlapping genes between the TWAS for AD and VaD, we identified genes associated with general cognitive function that were not explicitly identified by Davies et al. (2018)(21) which were associated with pre-clinical AD and VaD risk factors including cardiovascular diseases, immunity and Alzheimer’s neuropathology. Our TWAS also identified genes previously associated with cognitive domains, neuropathology, and psychiatric illness including reading-related skills and neural structures (*SEMA6D*(100) and *SETBP1*(101)), working memory tasks (*CDH13*(102)) and Schizophrenia (*HP*(103,104)*, C18orf1*(105) and *TMEM180*(106)). There are likely also distinct transcriptomic mechanisms that differentiate cognitive function and normal age-related brain changes from pathways related to dementia. Individuals who never develop dementia or significant cognitive decline still experience brain deterioration in normal aging that includes gray and white matter loss and ventricular enlargement which is accompanied by memory decline(107). Further, previous GWAS for general cognitive function and AD have shown few overlapping loci(21,108). In addition, studies of older individuals who are cognitively “resilient” with intact cognitive function, despite the presence of AD neuropathology, have found the genetic architecture of cognitive resilience to be distinct from that of AD(109). As such, relatively little is known about the pathways underlying cognitive aging in those without dementia. Thus, studying transcriptomic mechanisms that affect general cognitive function before development of dementia may shed light on cognitive aging without dementia.

We also compared genes identified by METRO after fine-mapping with those identified by TWAS-Fusion in Sargurupremraj et al. (2020)(22) and Bellenguez et al. (2022)(23). Among the 92 genes associated with WMH in Sargurupremraj et al. (2020)(22) and 23 genes identified by METRO, 10 genes overlapped. We note that the Sargurupremraj et al. (2020)(22) did not perform fine-mapping of their TWAS results, which is likely why we identified substantially fewer genes. There were also 10 overlapping genes among the 66 genes associated with AD in Bellenguez et al. (2022)(23) and 69 genes identified by METRO. For both TWAS comparisons, a relatively small number of genes overlap likely due to differences in eQTL prediction modeling. Sargurupremraj et al. (2020)(22) and Bellenguez et al. (2022)(23) used eQTL data from brain tissue, while we used eQTL data from transformed beta lymphocytes in blood tissue. While brain tissue is more relevant to WMH and AD phenotypes, blood cells do touch every cell bed that affects the brain, and are related to chronic inflammation, immunity, and oxidative stress, which are linked to cognitive performance and dementia. TWAS results from blood tissue in multiple ancestries provide complementary information to those reported in the GWAS.

Several limitations in the present study should be noted. First, our gene expression levels were measured using transformed B-lymphocytes from immortalized cell lines in GENOA. While transformed B-lymphocytes are a convenient source of DNA from blood tissue, we lack eQTL data for tissues that may be most relevant for AD and WMH (e.g., brain tissue, small brain vessels, and microglia). However, B-lymphocytes provide a unique and efficient way to examine the functional effects of genetic variations on gene expression that minimizes environmental influences(110). Second, METRO follows the standard TWAS approach of analyzing one gene at a time. Since genes residing in the same genomic region may share eQTLs or contain eQTL SNPs that are in LD with each other, the TWAS test statistics of genes in the same region may be highly correlated. To that end, it may be challenging to identify the truly biologically relevant genes among them(37,111). As such, we paired METRO with FOCUS to allow us to narrow down the list of potential causal genes for AD, VaD, and cognitive decline(37,112). Lastly, we primarily utilized EA GWAS that were publicly available with large sample sizes for general cognitive function, WMH, and AD. As expected, the gene expression prediction models constructed in the same ancestry as the GWAS (EA) tended to have larger contribution weights than AA. While we conducted a TWAS of AD in AA, the sample size of the AA GWAS likely did not allow us to properly power our TWAS. As such, a future direction would be to conduct TWAS of these traits using summary statistics from well-powered GWAS with AA ancestry or multiple ancestries as they become available.

Our study also has notable strengths. To our knowledge, our study is the first TWAS using expression mapping studies in multiple ancestries (EA and AA) to identify genes associated with cognitive function and neurocognitive disorders. By leveraging the complementary information in gene expression prediction models constructed in EA and AA, as well as the uncertainty in SNP prediction weights, we were able to conduct a highly powered TWAS to identify important gene-trait associations and transcriptomic mechanisms related to innate immunity, vascular dysfunction and neuroinflammation underlying AD, VaD, and general cognitive function. Using METRO, we were also able to estimate the ancestry contribution weights for specific genes and identify the extent to which a gene in EA or AA may contribute to the trait. However, it is noteworthy that the larger the contribution of the expression prediction models in the same ancestry as the GWAS (primarily EA, in this study) may allow for better predictive performance in the same ancestry. We also conducted FOCUS fine-mapping to narrow in on a list of putatively causal genes among multiple significant genes in a region. Our results suggest that there are similar pathways that contribute to healthy cognitive aging and progression of dementia, as well as distinct pathways that are unique to each neuropathology. By understanding overlapping and unique genes and transcriptomic mechanisms associated with each outcome, we may identify possible targets for prevention and/or treatments for cognitive aging and dementia.

## Conclusion

In the present study, we conducted a multi-ancestry TWAS in EA and AA to identify genes associated with general cognitive function, WMH and AD. We identified genes associated with innate immunity, vascular dysfunction, and neuroinflammation. The WMH and AD TWAS also indicated that downregulation of *ICA1L* may contribute to overlapping AD and VaD neuropathology. To our knowledge, this study is the first TWAS analysis using expression mapping studies in multiple ancestries to identify genes associated with cognitive function and neurocognitive disorders, which may help to identify gene targets for pharmaceutical or preventative treatment for dementia.

## Author Contributions

Conceptualization, D.L.C., J.A.S.; methodology, D.L.C., J.A.S., X.Z.; software, formal analysis, and investigation, D.L.C., Z.L., L.S., S.M.R.; resources, T.H.M. and S.L.R.K.; writing—original draft preparation, D.L.C., J.A.S.; writing—review and editing, D.L.C., S.M.R., W.Z., T.H.M., S.L.R.K., X.Z., J.A.S.; visualization, D.L.C., Z.L.; supervision, J.A.S.; funding acquisition, S.L.R.K. and J.A.S. All authors have read and agreed to the published version of the manuscript.

## Informed Consent Statement

Written informed consent was obtained from all subjects involved in the study.

## Data Availability Statement

The phenotype data and *APOE* genotypes used in the current study are available upon reasonable request to J.A.S. and S.L.R.K., and with a completed data use agreement (DUA). All other genotype data are available from the Database of Genotypes and Phenotypes (dbGaP): phs001401.v2.p1. Methylation and gene expression data are available from the Gene Expression Omnibus (GEO): GSE210256 and GSE138914. Due to IRB restriction, mapping of the sample IDs between genotype data (dbGaP) and methylation data (GEO) cannot be provided publicly, but is available upon written request to J.A.S. and S.L.R.K.

## Acknowledgments

The authors wish to thank the staff and participants of the GENOA study.

