## Supplementary Figures for "Multi-ancestry transcriptome-wide association studies of cognitive function, white matter hyperintensity, and Alzheimer’s disease"

**Table S1. Genes associated with general cognitive function using METRO followed by fine-mapping with FOCUS (N=266 genes; P<2.9x10^-6^)**

| **Gene** | **ENSG** | **alpha** | **w1** | **w2** | **P value** | **chr** | **Start** | **End** |
| --- | --- | --- | --- | --- | --- | --- | --- | --- |
| RNF123 | ENSG00000164068 | -0.23 | 0.58 | 0.42 | 3.20E-22 | 3 | 49726971 | 49758962 |
| RABEP2 | ENSG00000177548 | -1.16 | 0.68 | 0.32 | 4.20E-22 | 16 | 28915742 | 28947847 |
| GMPPB | ENSG00000173540 | -0.28 | 1.00 | 0.00 | 8.35E-22 | 3 | 49754277 | 49761406 |
| MST1 | ENSG00000173531 | -0.33 | 0.00 | 1.00 | 1.29E-21 | 3 | 49721380 | 49726934 |
| APEH | ENSG00000164062 | 0.95 | 0.00 | 1.00 | 2.30E-21 | 3 | 49711447 | 49721404 |
| IP6K1 | ENSG00000176095 | -0.76 | 0.62 | 0.38 | 4.32E-21 | 3 | 49761727 | 49823975 |
| UBA7 | ENSG00000182179 | 0.11 | 0.76 | 0.24 | 1.92E-20 | 3 | 49842642 | 49851386 |
| TUFM | ENSG00000178952 | -0.24 | 0.89 | 0.11 | 3.85E-20 | 16 | 28853732 | 28857669 |
| FOXO6 | ENSG00000204060 | 1.15 | 0.87 | 0.13 | 8.29E-20 | 1 | 41827594 | 41849262 |
| NFKB2 | ENSG00000077150 | 0.30 | 0.00 | 1.00 | 3.67E-19 | 10 | 104153867 | 104162281 |
| PSD | ENSG00000059915 | 0.21 | 0.00 | 1.00 | 4.16E-19 | 10 | 104162374 | 104181296 |
| STAU1 | ENSG00000124214 | 0.17 | 0.52 | 0.48 | 9.53E-19 | 20 | 47729876 | 47804904 |
| CSE1L | ENSG00000124207 | -0.55 | 0.61 | 0.39 | 1.74E-18 | 20 | 47662783 | 47713497 |
| ARFGEF2 | ENSG00000124198 | 0.11 | 1.00 | 0.00 | 3.20E-18 | 20 | 47538248 | 47653230 |
| USP4 | ENSG00000114316 | -0.76 | 0.27 | 0.73 | 4.14E-18 | 3 | 49314577 | 49378145 |
| MEF2C | ENSG00000081189 | -0.26 | 0.57 | 0.43 | 2.32E-17 | 5 | 88012934 | 88200074 |
| PTPRD | ENSG00000153707 | -0.70 | 1.00 | 0.00 | 1.03E-15 | 9 | 8314246 | 10613002 |
| NEGR1 | ENSG00000172260 | -0.28 | 0.22 | 0.78 | 1.23E-15 | 1 | 71861626 | 72748222 |
| SEMA3G | ENSG00000010319 | -2.55 | 0.00 | 1.00 | 1.96E-15 | 3 | 52467051 | 52479119 |
| ABT1 | ENSG00000146109 | 0.61 | 0.39 | 0.61 | 2.24E-15 | 6 | 26597181 | 26600967 |
| FOXP1 | ENSG00000114861 | 1.10 | 0.93 | 0.07 | 3.34E-15 | 3 | 71003844 | 71633129 |
| TET2 | ENSG00000168769 | -0.12 | 0.83 | 0.17 | 3.55E-15 | 4 | 106067032 | 106200973 |
| ZNF322 | ENSG00000181315 | 0.33 | 0.10 | 0.90 | 7.88E-15 | 6 | 26634611 | 26659980 |
| HMGN4 | ENSG00000182952 | -0.09 | 0.00 | 1.00 | 1.08E-14 | 6 | 26538594 | 26547161 |
| NISCH | ENSG00000010322 | -0.25 | 1.00 | 0.00 | 5.24E-14 | 3 | 52489134 | 52527084 |
| RBFOX1 | ENSG00000078328 | 0.22 | 0.96 | 0.04 | 8.64E-14 | 16 | 5289803 | 7763342 |
| ELAVL2 | ENSG00000107105 | -0.56 | 0.00 | 1.00 | 1.43E-13 | 9 | 23690102 | 23826335 |
| IL27 | ENSG00000197272 | 1.73 | 1.00 | 0.00 | 1.95E-13 | 16 | 28510683 | 28523372 |
| TSNARE1 | ENSG00000171045 | -0.20 | 0.00 | 1.00 | 4.54E-13 | 8 | 143293441 | 143484543 |
| KCNJ3 | ENSG00000162989 | 0.34 | 0.00 | 1.00 | 5.53E-13 | 2 | 155554367 | 155714866 |
| MTMR4 | ENSG00000108389 | -0.12 | 0.00 | 1.00 | 6.56E-13 | 17 | 56566890 | 56595266 |
| AFF3 | ENSG00000144218 | -0.31 | 0.00 | 1.00 | 1.32E-12 | 2 | 100161881 | 100808890 |
| LACE1 | ENSG00000135537 | -0.24 | 0.27 | 0.73 | 1.42E-12 | 6 | 108616195 | 108847999 |
| ATXN1 | ENSG00000124788 | 0.82 | 0.00 | 1.00 | 1.83E-12 | 6 | 16299343 | 16761722 |
| PEF1 | ENSG00000162517 | 0.11 | 0.79 | 0.21 | 2.25E-12 | 1 | 32095467 | 32110497 |
| LONRF2 | ENSG00000170500 | 0.45 | 0.00 | 1.00 | 3.21E-12 | 2 | 100888337 | 100938963 |
| OR2J1 | ENSG00000204702 | -0.12 | 1.00 | 0.00 | 3.23E-12 | 6 | 29067267 | 29070478 |
| HSF5 | ENSG00000176160 | -0.73 | 0.60 | 0.40 | 3.93E-12 | 17 | 56497528 | 56565769 |
| ST3GAL3 | ENSG00000126091 | -0.09 | 0.90 | 0.10 | 5.88E-12 | 1 | 44171495 | 44396837 |
| NKIRAS1 | ENSG00000197885 | -0.08 | 1.00 | 0.00 | 6.15E-12 | 3 | 23931442 | 23988082 |
| COL16A1 | ENSG00000084636 | -1.07 | 0.00 | 1.00 | 6.99E-12 | 1 | 32117864 | 32169920 |
| DCC | ENSG00000187323 | -2.90 | 0.00 | 1.00 | 7.58E-12 | 18 | 49866567 | 51062273 |
| FOXO3 | ENSG00000118689 | -1.70 | 0.00 | 1.00 | 1.09E-11 | 6 | 108881038 | 109005977 |
| PRSS16 | ENSG00000112812 | 0.30 | 0.53 | 0.47 | 1.11E-11 | 6 | 27215480 | 27224403 |
| 4-SEP | ENSG00000108387 | -0.60 | 0.00 | 1.00 | 1.49E-11 | 17 | 56597611 | 56621729 |
| QRICH1 | ENSG00000198218 | 0.09 | 0.00 | 1.00 | 1.73E-11 | 3 | 49067140 | 49131796 |
| RNF43 | ENSG00000108375 | -0.21 | 0.00 | 1.00 | 1.82E-11 | 17 | 56431037 | 56494956 |
| ZNF193 | ENSG00000137185 | 0.25 | 0.41 | 0.59 | 1.84E-11 | 6 | 28192664 | 28201265 |
| ARF5 | ENSG00000004059 | 0.15 | 1.00 | 0.00 | 2.40E-11 | 7 | 127228440 | 127231754 |
| ZNF184 | ENSG00000096654 | 0.21 | 1.00 | 0.00 | 2.54E-11 | 6 | 27418522 | 27440897 |
| DPP4 | ENSG00000197635 | -1.30 | 0.02 | 0.98 | 2.70E-11 | 2 | 162848755 | 162930904 |
| OR2J3 | ENSG00000204701 | -0.29 | 0.00 | 1.00 | 3.13E-11 | 6 | 29075835 | 29082547 |
| SP4 | ENSG00000105866 | 0.09 | 1.00 | 0.00 | 3.98E-11 | 7 | 21467661 | 21554440 |
| FSCN3 | ENSG00000106328 | 0.51 | 0.90 | 0.10 | 4.04E-11 | 7 | 127231463 | 127242198 |
| GCC1 | ENSG00000179562 | 0.13 | 0.88 | 0.12 | 4.72E-11 | 7 | 127220682 | 127233665 |
| OR2H1 | ENSG00000204688 | -0.23 | 0.00 | 1.00 | 4.98E-11 | 6 | 29424932 | 29432105 |
| SGCZ | ENSG00000185053 | 0.33 | 1.00 | 0.00 | 5.00E-11 | 8 | 13942354 | 15095940 |
| FBXO41 | ENSG00000163013 | -0.21 | 0.99 | 0.01 | 5.10E-11 | 2 | 73481810 | 73511606 |
| NR1D2 | ENSG00000174738 | -0.17 | 0.00 | 1.00 | 5.30E-11 | 3 | 23986777 | 24022108 |
| DHODH | ENSG00000102967 | -0.15 | 0.90 | 0.10 | 5.44E-11 | 16 | 72042487 | 72061563 |
| HP | ENSG00000257017 | -1.24 | 0.19 | 0.81 | 7.42E-11 | 16 | 72088404 | 72094954 |
| ATF4 | ENSG00000128272 | -0.48 | 0.09 | 0.91 | 7.66E-11 | 22 | 39915700 | 39918688 |
| TRIM27 | ENSG00000204713 | -0.92 | 0.00 | 1.00 | 9.36E-11 | 6 | 28870779 | 28891765 |
| HIST1H2AG | ENSG00000196787 | -0.17 | 0.00 | 1.00 | 1.00E-10 | 6 | 27100822 | 27101314 |
| CCT7 | ENSG00000135624 | 0.15 | 0.00 | 1.00 | 1.01E-10 | 2 | 73460548 | 73480149 |
| THRB | ENSG00000151090 | 0.48 | 0.00 | 1.00 | 1.37E-10 | 3 | 24158644 | 24537247 |
| ZKSCAN4 | ENSG00000187626 | 0.42 | 0.23 | 0.77 | 1.44E-10 | 6 | 28209475 | 28220047 |
| SND1 | ENSG00000197157 | -1.77 | 0.01 | 0.99 | 1.51E-10 | 7 | 127292248 | 127732661 |
| PURA | ENSG00000185129 | 0.56 | 0.94 | 0.06 | 1.65E-10 | 5 | 139487362 | 139505204 |
| SLC6A9 | ENSG00000196517 | 0.25 | 0.37 | 0.63 | 1.71E-10 | 1 | 44457172 | 44497139 |
| MGAT3 | ENSG00000128268 | 0.38 | 0.00 | 1.00 | 1.91E-10 | 22 | 39853017 | 39888199 |
| POU6F2 | ENSG00000106536 | 0.55 | 0.00 | 1.00 | 1.92E-10 | 7 | 39017509 | 39532694 |
| PPM1M | ENSG00000164088 | 0.12 | 0.58 | 0.42 | 2.21E-10 | 3 | 52279775 | 52284615 |
| NPAS3 | ENSG00000151322 | 0.72 | 1.00 | 0.00 | 2.44E-10 | 14 | 33403602 | 34290069 |
| IST1 | ENSG00000182149 | 0.17 | 0.00 | 1.00 | 2.75E-10 | 16 | 71879899 | 71965102 |
| PRKAG1 | ENSG00000181929 | 0.11 | 1.00 | 0.00 | 2.76E-10 | 12 | 49396057 | 49412590 |
| HIST1H2BL | ENSG00000185130 | -0.05 | 0.03 | 0.97 | 2.77E-10 | 6 | 27775257 | 27775707 |
| PRADC1 | ENSG00000135617 | 0.32 | 0.63 | 0.37 | 2.79E-10 | 2 | 73455138 | 73460367 |
| CYSTM1 | ENSG00000120306 | 0.35 | 0.00 | 1.00 | 2.82E-10 | 5 | 139554741 | 139661637 |
| IPO9 | ENSG00000198700 | 0.09 | 0.00 | 1.00 | 2.92E-10 | 1 | 201798277 | 201853419 |
| EGR4 | ENSG00000135625 | 0.95 | 0.87 | 0.13 | 3.14E-10 | 2 | 73518057 | 73520829 |
| PDE4C | ENSG00000105650 | -0.40 | 0.00 | 1.00 | 3.23E-10 | 19 | 18319462 | 18359010 |
| HBEGF | ENSG00000113070 | -0.50 | 0.00 | 1.00 | 3.38E-10 | 5 | 139712428 | 139726188 |
| KIAA1683 | ENSG00000130518 | -0.19 | 0.00 | 1.00 | 3.52E-10 | 19 | 18367908 | 18385310 |
| HPR | ENSG00000261701 | -0.62 | 0.02 | 0.98 | 3.79E-10 | 16 | 72097047 | 72111145 |
| NKAIN2 | ENSG00000188580 | 24.44 | 0.00 | 1.00 | 5.73E-10 | 6 | 124125010 | 125146786 |
| RNF39 | ENSG00000204618 | -0.53 | 0.66 | 0.34 | 5.95E-10 | 6 | 30038043 | 30043626 |
| SRPK2 | ENSG00000135250 | -0.05 | 1.00 | 0.00 | 6.18E-10 | 7 | 104751151 | 105039755 |
| SFXN5 | ENSG00000144040 | 0.07 | 1.00 | 0.00 | 6.30E-10 | 2 | 73169165 | 73302747 |
| MLL5 | ENSG00000005483 | 0.25 | 1.00 | 0.00 | 6.40E-10 | 7 | 104581390 | 104755466 |
| RBL2 | ENSG00000103479 | -0.11 | 0.00 | 1.00 | 6.89E-10 | 16 | 53467889 | 53525560 |
| PFDN1 | ENSG00000113068 | 0.16 | 0.76 | 0.24 | 7.66E-10 | 5 | 139624620 | 139682698 |
| HIST1H2BJ | ENSG00000124635 | 0.04 | 1.00 | 0.00 | 7.95E-10 | 6 | 27093676 | 27100574 |
| SPPL2C | ENSG00000185294 | -0.82 | 0.41 | 0.59 | 8.29E-10 | 17 | 43922247 | 43924433 |
| CDH8 | ENSG00000150394 | 0.37 | 0.67 | 0.33 | 8.67E-10 | 16 | 61681146 | 62070939 |
| FAM109B | ENSG00000177096 | -0.13 | 1.00 | 0.00 | 1.03E-09 | 22 | 42470252 | 42475442 |
| PTPRO | ENSG00000151490 | -0.19 | 0.00 | 1.00 | 1.08E-09 | 12 | 15475191 | 15755109 |
| SLC39A8 | ENSG00000138821 | 0.18 | 0.00 | 1.00 | 1.35E-09 | 4 | 103172237 | 103352415 |
| TNFRSF13C | ENSG00000159958 | -0.01 | 0.00 | 1.00 | 1.37E-09 | 22 | 42318036 | 42322810 |
| CPXM2 | ENSG00000121898 | -0.87 | 0.30 | 0.70 | 1.49E-09 | 10 | 125465723 | 125699783 |
| PLCL1 | ENSG00000115896 | 0.45 | 0.00 | 1.00 | 1.60E-09 | 2 | 198669317 | 199437305 |
| NCOA2 | ENSG00000140396 | 0.32 | 0.09 | 0.91 | 2.03E-09 | 8 | 71022017 | 71316043 |
| PKD2L1 | ENSG00000107593 | 0.26 | 0.70 | 0.30 | 2.28E-09 | 10 | 102047906 | 102090021 |
| CDKAL1 | ENSG00000145996 | 0.63 | 0.00 | 1.00 | 2.31E-09 | 6 | 20534688 | 21232635 |
| SORT1 | ENSG00000134243 | -0.10 | 0.72 | 0.28 | 2.36E-09 | 1 | 109852190 | 109940540 |
| TANK | ENSG00000136560 | -0.43 | 0.69 | 0.31 | 2.37E-09 | 2 | 161993419 | 162092741 |
| SUOX | ENSG00000139531 | -0.08 | 0.57 | 0.43 | 2.38E-09 | 12 | 56390964 | 56400425 |
| MLL2 | ENSG00000167548 | 0.23 | 0.44 | 0.56 | 3.06E-09 | 12 | 49412758 | 49454577 |
| LSM4 | ENSG00000130520 | 0.43 | 0.55 | 0.45 | 4.06E-09 | 19 | 18417046 | 18433922 |
| TIMM17A | ENSG00000134375 | 0.38 | 0.14 | 0.86 | 4.99E-09 | 1 | 201924631 | 201939792 |
| RHEBL1 | ENSG00000167550 | -1.80 | 0.14 | 0.86 | 5.03E-09 | 12 | 49458459 | 49463808 |
| CALN1 | ENSG00000183166 | -0.67 | 0.43 | 0.57 | 5.38E-09 | 7 | 71244476 | 71912136 |
| PDE4D | ENSG00000113448 | -0.19 | 1.00 | 0.00 | 6.24E-09 | 5 | 58264865 | 59817947 |
| DDN | ENSG00000181418 | -0.28 | 1.00 | 0.00 | 6.81E-09 | 12 | 49388932 | 49393158 |
| CWF19L1 | ENSG00000095485 | 0.07 | 0.00 | 1.00 | 7.48E-09 | 10 | 101992055 | 102027437 |
| NKX2-1 | ENSG00000136352 | -0.42 | 0.56 | 0.44 | 8.23E-09 | 14 | 36985597 | 36990354 |
| GDF15 | ENSG00000130513 | 0.10 | 0.00 | 1.00 | 9.47E-09 | 19 | 18485541 | 18499986 |
| NFIX | ENSG00000008441 | 0.24 | 0.03 | 0.97 | 1.02E-08 | 19 | 13106289 | 13209610 |
| SNX29 | ENSG00000048471 | 0.16 | 0.00 | 1.00 | 1.03E-08 | 16 | 12070591 | 12668144 |
| AUTS2 | ENSG00000158321 | -0.25 | 0.00 | 1.00 | 1.16E-08 | 7 | 69063282 | 70258492 |
| CHUK | ENSG00000213341 | 0.34 | 0.00 | 1.00 | 1.21E-08 | 10 | 101948057 | 101989353 |
| RALYL | ENSG00000184672 | -0.34 | 0.00 | 1.00 | 1.66E-08 | 8 | 85095022 | 85834079 |
| PSMA5 | ENSG00000143106 | 0.00 | 0.50 | 0.50 | 1.87E-08 | 1 | 109941664 | 109969070 |
| PRKAR2B | ENSG00000005249 | 0.41 | 0.45 | 0.55 | 1.93E-08 | 7 | 106685150 | 106802256 |
| EPS8 | ENSG00000151491 | -1.55 | 0.33 | 0.67 | 2.10E-08 | 12 | 15773068 | 16035263 |
| LYL1 | ENSG00000104903 | 0.25 | 0.00 | 1.00 | 2.25E-08 | 19 | 13209847 | 13213975 |
| PSMC3 | ENSG00000165916 | -2.36 | 0.04 | 0.96 | 2.29E-08 | 11 | 47440320 | 47448024 |
| LRRC14 | ENSG00000160959 | 0.11 | 0.00 | 1.00 | 2.34E-08 | 8 | 145743376 | 145750556 |
| WNT10B | ENSG00000169884 | 0.42 | 0.00 | 1.00 | 2.49E-08 | 12 | 49359123 | 49365518 |
| RUNX1T1 | ENSG00000079102 | 0.42 | 0.94 | 0.06 | 2.51E-08 | 8 | 92967195 | 93115514 |
| PET112 | ENSG00000059691 | -0.05 | 0.90 | 0.10 | 3.00E-08 | 4 | 152591656 | 152682159 |
| NKX2-8 | ENSG00000136327 | -0.20 | 0.00 | 1.00 | 3.10E-08 | 14 | 37049209 | 37051819 |
| GLYCTK | ENSG00000168237 | -0.13 | 0.67 | 0.33 | 3.20E-08 | 3 | 52321105 | 52329273 |
| CAMK2N1 | ENSG00000162545 | 0.18 | 0.59 | 0.41 | 3.50E-08 | 1 | 20808884 | 20812703 |
| CKB | ENSG00000166165 | 0.10 | 0.12 | 0.88 | 3.68E-08 | 14 | 103986004 | 103989170 |
| MYLK | ENSG00000065534 | 0.47 | 0.47 | 0.53 | 3.71E-08 | 3 | 123328896 | 123603179 |
| CALR | ENSG00000179218 | 0.76 | 0.00 | 1.00 | 3.80E-08 | 19 | 13049392 | 13055303 |
| JMJD1C | ENSG00000171988 | 0.30 | 0.31 | 0.69 | 4.49E-08 | 10 | 64926981 | 65281610 |
| LRRC25 | ENSG00000175489 | 0.13 | 1.00 | 0.00 | 4.61E-08 | 19 | 18501947 | 18508432 |
| PPP1R16A | ENSG00000160972 | 0.31 | 0.45 | 0.55 | 4.66E-08 | 8 | 145703352 | 145727504 |
| KCNJ6 | ENSG00000157542 | 0.28 | 0.93 | 0.07 | 5.22E-08 | 21 | 38979675 | 39493439 |
| EIF2B5 | ENSG00000145191 | 0.96 | 0.45 | 0.55 | 5.93E-08 | 3 | 183852826 | 183863915 |
| GADD45GIP1 | ENSG00000179271 | -0.41 | 1.00 | 0.00 | 5.97E-08 | 19 | 13063933 | 13068037 |
| TONSL | ENSG00000160949 | 0.40 | 0.48 | 0.52 | 6.17E-08 | 8 | 145654158 | 145669823 |
| ELK4 | ENSG00000158711 | -0.33 | 1.00 | 0.00 | 6.92E-08 | 1 | 205566684 | 205601139 |
| NMNAT2 | ENSG00000157064 | 1.60 | 0.76 | 0.24 | 7.81E-08 | 1 | 183217372 | 183387515 |
| RAD23A | ENSG00000179262 | 0.16 | 1.00 | 0.00 | 7.92E-08 | 19 | 13056669 | 13064456 |
| MACROD2 | ENSG00000172264 | -0.12 | 1.00 | 0.00 | 8.80E-08 | 20 | 13976015 | 16033842 |
| DBN1 | ENSG00000113758 | 0.07 | 1.00 | 0.00 | 9.91E-08 | 5 | 176883609 | 176901402 |
| FAM193A | ENSG00000125386 | 0.09 | 0.10 | 0.90 | 1.04E-07 | 4 | 2538374 | 2734300 |
| FARSA | ENSG00000179115 | 0.11 | 1.00 | 0.00 | 1.13E-07 | 19 | 13033293 | 13044851 |
| NRBF2 | ENSG00000148572 | 0.25 | 0.80 | 0.20 | 1.16E-07 | 10 | 64893007 | 64914791 |
| ZSWIM6 | ENSG00000130449 | -1.09 | 0.00 | 1.00 | 1.18E-07 | 5 | 60628085 | 60841999 |
| CCDC14 | ENSG00000175455 | -0.13 | 0.00 | 1.00 | 1.28E-07 | 3 | 123616152 | 123680255 |
| CDH13 | ENSG00000140945 | -0.67 | 1.00 | 0.00 | 1.29E-07 | 16 | 82660570 | 83834245 |
| MFSD4 | ENSG00000174514 | -0.07 | 1.00 | 0.00 | 1.31E-07 | 1 | 205538013 | 205572046 |
| EPHA5 | ENSG00000145242 | 0.29 | 0.00 | 1.00 | 1.53E-07 | 4 | 66185281 | 66536213 |
| TSHZ3 | ENSG00000121297 | -0.79 | 0.55 | 0.45 | 1.54E-07 | 19 | 31640885 | 31840342 |
| SLC39A4 | ENSG00000147804 | -1.75 | 0.00 | 1.00 | 1.65E-07 | 8 | 145635126 | 145642228 |
| DAND5 | ENSG00000179284 | 1.23 | 0.00 | 1.00 | 1.66E-07 | 19 | 13075973 | 13085574 |
| PDCL3 | ENSG00000115539 | -0.11 | 1.00 | 0.00 | 1.79E-07 | 2 | 101179455 | 101193201 |
| DCAF11 | ENSG00000100897 | -0.77 | 0.03 | 0.97 | 1.84E-07 | 14 | 24583404 | 24594451 |
| GCDH | ENSG00000105607 | 0.18 | 1.00 | 0.00 | 1.96E-07 | 19 | 13001974 | 13025021 |
| TMEM180 | ENSG00000138111 | -0.09 | 1.00 | 0.00 | 2.03E-07 | 10 | 104221152 | 104236802 |
| AGAP1 | ENSG00000157985 | -0.62 | 0.33 | 0.67 | 2.35E-07 | 2 | 236402687 | 237040444 |
| KLF1 | ENSG00000105610 | -1.72 | 0.18 | 0.82 | 2.49E-07 | 19 | 12995236 | 12998015 |
| CDH4 | ENSG00000179242 | 0.36 | 0.51 | 0.49 | 2.63E-07 | 20 | 59827317 | 60515673 |
| CFB | ENSG00000243649 | -1.58 | 0.93 | 0.07 | 2.64E-07 | 6 | 31913427 | 31919861 |
| MAML2 | ENSG00000184384 | 0.24 | 0.00 | 1.00 | 2.71E-07 | 11 | 95709762 | 96076359 |
| IL34 | ENSG00000157368 | 0.44 | 0.86 | 0.14 | 2.75E-07 | 16 | 70613798 | 70694585 |
| CPEB1 | ENSG00000214575 | 0.12 | 0.00 | 1.00 | 2.90E-07 | 15 | 83211951 | 83317612 |
| MTSS1L | ENSG00000132613 | 0.48 | 1.00 | 0.00 | 2.95E-07 | 16 | 70695107 | 70719956 |
| GRK6 | ENSG00000198055 | -0.23 | 0.67 | 0.33 | 3.02E-07 | 5 | 176830205 | 176869902 |
| IL17D | ENSG00000172458 | 0.41 | 0.50 | 0.50 | 3.23E-07 | 13 | 21276266 | 21297237 |
| FMNL3 | ENSG00000161791 | -0.26 | 0.44 | 0.56 | 3.28E-07 | 12 | 50030282 | 50101948 |
| RORA | ENSG00000069667 | 0.25 | 0.01 | 0.99 | 3.44E-07 | 15 | 60780483 | 61521501 |
| FBXL17 | ENSG00000145743 | 0.37 | 0.77 | 0.23 | 3.45E-07 | 5 | 107194736 | 107717799 |
| NCAM1 | ENSG00000149294 | -0.89 | 0.72 | 0.28 | 3.58E-07 | 11 | 112831969 | 113149158 |
| CSRNP3 | ENSG00000178662 | 0.34 | 0.53 | 0.47 | 3.60E-07 | 2 | 166326157 | 166545917 |
| SSBP2 | ENSG00000145687 | 0.42 | 0.63 | 0.37 | 3.82E-07 | 5 | 80708623 | 81047616 |
| CPNE6 | ENSG00000100884 | 0.26 | 1.00 | 0.00 | 4.05E-07 | 14 | 24540046 | 24547309 |
| FITM1 | ENSG00000139914 | -0.42 | 0.00 | 1.00 | 4.07E-07 | 14 | 24599868 | 24602058 |
| SEMA3E | ENSG00000170381 | 0.20 | 0.16 | 0.84 | 4.08E-07 | 7 | 82992554 | 83278455 |
| NDUFAF2 | ENSG00000164182 | -0.17 | 1.00 | 0.00 | 4.27E-07 | 5 | 60241004 | 60450358 |
| LDB2 | ENSG00000169744 | 0.23 | 0.00 | 1.00 | 4.45E-07 | 4 | 16503164 | 16900301 |
| MGST2 | ENSG00000085871 | 0.07 | 0.76 | 0.24 | 4.92E-07 | 4 | 140586922 | 140661899 |
| RNF4 | ENSG00000063978 | 0.05 | 0.29 | 0.71 | 5.04E-07 | 4 | 2463947 | 2627047 |
| RAI1 | ENSG00000108557 | -0.08 | 0.00 | 1.00 | 5.43E-07 | 17 | 17584772 | 17714767 |
| IGSF9B | ENSG00000080854 | 0.26 | 1.00 | 0.00 | 5.50E-07 | 11 | 133766333 | 133826863 |
| ANKS1B | ENSG00000185046 | -0.26 | 0.70 | 0.30 | 5.51E-07 | 12 | 99120235 | 100378714 |
| C6orf108 | ENSG00000112667 | -0.05 | 0.75 | 0.25 | 5.66E-07 | 6 | 43193367 | 43197219 |
| C6orf25 | ENSG00000204420 | -0.18 | 0.20 | 0.80 | 5.80E-07 | 6 | 31686371 | 31694491 |
| CDH9 | ENSG00000113100 | -0.28 | 1.00 | 0.00 | 6.16E-07 | 5 | 26880706 | 27121257 |
| TNRC6A | ENSG00000090905 | -0.14 | 1.00 | 0.00 | 6.19E-07 | 16 | 24621530 | 24838953 |
| CRAT | ENSG00000095321 | -0.03 | 0.00 | 1.00 | 6.32E-07 | 9 | 131856421 | 131873468 |
| C18orf1 | ENSG00000168675 | 0.69 | 1.00 | 0.00 | 6.32E-07 | 18 | 13217497 | 13652754 |
| C17orf59 | ENSG00000196544 | -0.35 | 0.55 | 0.45 | 6.45E-07 | 17 | 8091663 | 8093498 |
| SLC34A1 | ENSG00000131183 | -0.33 | 1.00 | 0.00 | 6.89E-07 | 5 | 176806236 | 176825849 |
| CEP192 | ENSG00000101639 | 0.03 | 0.10 | 0.90 | 6.97E-07 | 18 | 12991361 | 13125051 |
| FBXL4 | ENSG00000112234 | -0.11 | 1.00 | 0.00 | 7.02E-07 | 6 | 99316411 | 99395882 |
| NFAM1 | ENSG00000235568 | 0.17 | 0.00 | 1.00 | 7.09E-07 | 22 | 42776413 | 42828409 |
| C20orf173 | ENSG00000125975 | -0.19 | 0.70 | 0.30 | 7.20E-07 | 20 | 34111014 | 34117481 |
| PRR7 | ENSG00000131188 | 0.51 | 0.00 | 1.00 | 7.29E-07 | 5 | 176873446 | 176883287 |
| SCN2A | ENSG00000136531 | 0.14 | 0.00 | 1.00 | 8.00E-07 | 2 | 166051503 | 166248820 |
| SP2 | ENSG00000167182 | 0.16 | 0.00 | 1.00 | 8.10E-07 | 17 | 45973516 | 46006323 |
| CPD | ENSG00000108582 | -0.08 | 0.55 | 0.45 | 8.37E-07 | 17 | 28705945 | 28797007 |
| CRIP3 | ENSG00000146215 | 0.31 | 1.00 | 0.00 | 8.49E-07 | 6 | 43267448 | 43276564 |
| SHISA9 | ENSG00000237515 | -0.53 | 0.28 | 0.72 | 8.58E-07 | 16 | 12995455 | 13334273 |
| SEMA6D | ENSG00000137872 | -0.76 | 1.00 | 0.00 | 8.84E-07 | 15 | 47476298 | 48066425 |
| XXbac-BPG32J3.19 | ENSG00000250641 | 0.27 | 0.71 | 0.29 | 8.98E-07 | 6 | 31674681 | 31685695 |
| MAML3 | ENSG00000196782 | 0.15 | 0.88 | 0.12 | 9.55E-07 | 4 | 140637907 | 141075338 |
| SMG7 | ENSG00000116698 | 0.00 | 0.50 | 0.50 | 9.78E-07 | 1 | 183441351 | 183567381 |
| SLC22A7 | ENSG00000137204 | -0.21 | 0.00 | 1.00 | 1.03E-06 | 6 | 43263432 | 43273276 |
| CADM2 | ENSG00000175161 | -0.11 | 0.82 | 0.18 | 1.04E-06 | 3 | 85008140 | 86123579 |
| PSME2 | ENSG00000100911 | -0.72 | 0.04 | 0.96 | 1.04E-06 | 14 | 24612571 | 24616779 |
| GPD2 | ENSG00000115159 | 0.20 | 1.00 | 0.00 | 1.13E-06 | 2 | 157291802 | 157470247 |
| PTPRT | ENSG00000196090 | -0.66 | 0.22 | 0.78 | 1.16E-06 | 20 | 40701392 | 41818610 |
| AKAP6 | ENSG00000151320 | 0.00 | 0.50 | 0.50 | 1.17E-06 | 14 | 32798504 | 33306890 |
| MTMR2 | ENSG00000087053 | 1.64 | 0.00 | 1.00 | 1.18E-06 | 11 | 95554930 | 95658479 |
| C19orf81 | ENSG00000235034 | -0.61 | 0.62 | 0.38 | 1.22E-06 | 19 | 51152702 | 51162567 |
| RMI1 | ENSG00000178966 | -0.28 | 0.63 | 0.37 | 1.28E-06 | 9 | 86595713 | 86618989 |
| CALML5 | ENSG00000178372 | 0.37 | 0.01 | 0.99 | 1.28E-06 | 10 | 5540660 | 5541533 |
| ATP5H | ENSG00000167863 | 6.63 | 0.02 | 0.98 | 1.28E-06 | 17 | 73034958 | 73043080 |
| TMBIM6 | ENSG00000139644 | 0.05 | 1.00 | 0.00 | 1.32E-06 | 12 | 50101508 | 50158717 |
| RGSL1 | ENSG00000121446 | -0.14 | 1.00 | 0.00 | 1.34E-06 | 1 | 182378327 | 182529734 |
| AVL9 | ENSG00000105778 | -0.08 | 0.00 | 1.00 | 1.38E-06 | 7 | 32535038 | 32628338 |
| TYW5 | ENSG00000162971 | 0.06 | 0.42 | 0.58 | 1.38E-06 | 2 | 200793636 | 200820459 |
| PSME1 | ENSG00000092010 | -1.04 | 0.00 | 1.00 | 1.49E-06 | 14 | 24605372 | 24608176 |
| KCNK3 | ENSG00000171303 | 1.06 | 0.36 | 0.64 | 1.51E-06 | 2 | 26915590 | 26956288 |
| HSPA1A | ENSG00000204389 | -0.48 | 0.06 | 0.94 | 1.65E-06 | 6 | 31783320 | 31785723 |
| LMF1 | ENSG00000103227 | -0.04 | 0.93 | 0.07 | 1.71E-06 | 16 | 903634 | 1031318 |
| MCRS1 | ENSG00000187778 | -0.04 | 1.00 | 0.00 | 1.76E-06 | 12 | 49950327 | 49961928 |
| SPAG4 | ENSG00000061656 | -0.12 | 0.25 | 0.75 | 1.82E-06 | 20 | 34203751 | 34209016 |
| SLC5A11 | ENSG00000158865 | 0.18 | 1.00 | 0.00 | 1.94E-06 | 16 | 24857162 | 24922949 |
| EXT1 | ENSG00000182197 | -0.09 | 0.00 | 1.00 | 1.94E-06 | 8 | 118806729 | 119124065 |
| MAST4 | ENSG00000069020 | 0.11 | 0.97 | 0.03 | 1.94E-06 | 5 | 65892208 | 66465421 |
| ROMO1 | ENSG00000125995 | -0.34 | 0.37 | 0.63 | 1.97E-06 | 20 | 34287194 | 34288906 |
| TMEM170B | ENSG00000205269 | -0.06 | 0.81 | 0.19 | 1.98E-06 | 6 | 11537982 | 11583757 |
| RPS17L | ENSG00000182774 | 0.23 | 0.83 | 0.17 | 2.00E-06 | 15 | 83205501 | 83209210 |
| LRRC9 | ENSG00000131951 | -0.14 | 0.00 | 1.00 | 2.01E-06 | 14 | 60386431 | 60530277 |
| ZNF638 | ENSG00000075292 | -0.11 | 0.00 | 1.00 | 2.04E-06 | 2 | 71503691 | 71662199 |
| C9orf64 | ENSG00000165118 | -0.07 | 1.00 | 0.00 | 2.05E-06 | 9 | 86553226 | 86571901 |
| SH3RF3 | ENSG00000172985 | 0.08 | 0.83 | 0.17 | 2.09E-06 | 2 | 109745661 | 110262211 |
| BCL11A | ENSG00000119866 | -0.28 | 0.58 | 0.42 | 2.11E-06 | 2 | 60677655 | 60781602 |
| MACROD1 | ENSG00000133315 | 0.21 | 1.00 | 0.00 | 2.14E-06 | 11 | 63766030 | 63933585 |
| PURG | ENSG00000172733 | 0.15 | 0.14 | 0.86 | 2.15E-06 | 8 | 30853318 | 30891231 |
| RP11-468E2.4 | ENSG00000259529 | -2.84 | 0.00 | 1.00 | 2.15E-06 | 14 | 24616757 | 24635661 |
| CUL9 | ENSG00000112659 | 0.20 | 0.29 | 0.71 | 2.21E-06 | 6 | 43149922 | 43192325 |
| LARGE | ENSG00000133424 | -0.18 | 0.92 | 0.08 | 2.21E-06 | 22 | 33558212 | 34318829 |
| VEGFA | ENSG00000112715 | 0.18 | 0.12 | 0.88 | 2.21E-06 | 6 | 43737921 | 43754224 |
| PHF20 | ENSG00000025293 | -0.10 | 0.99 | 0.01 | 2.25E-06 | 20 | 34359896 | 34538292 |
| SETBP1 | ENSG00000152217 | 0.25 | 0.89 | 0.11 | 2.25E-06 | 18 | 42260138 | 42648475 |
| C2orf47 | ENSG00000162972 | 0.31 | 0.48 | 0.52 | 2.29E-06 | 2 | 200820040 | 200873263 |
| KBTBD2 | ENSG00000170852 | -0.90 | 0.00 | 1.00 | 2.48E-06 | 7 | 32907784 | 32933743 |
| CNGB3 | ENSG00000170289 | 3.10 | 1.00 | 0.00 | 2.56E-06 | 8 | 87566205 | 87755903 |
| PCK2 | ENSG00000100889 | 0.35 | 0.00 | 1.00 | 2.56E-06 | 14 | 24563262 | 24579807 |
| DCAF5 | ENSG00000139990 | 0.16 | 1.00 | 0.00 | 2.64E-06 | 14 | 69517598 | 69619867 |
| PRPF38A | ENSG00000134748 | 0.20 | 0.17 | 0.83 | 2.67E-06 | 1 | 52870274 | 52886508 |
| SREBF1 | ENSG00000072310 | -0.35 | 0.41 | 0.59 | 2.67E-06 | 17 | 17713713 | 17740316 |
| NCKAP5L | ENSG00000167566 | 0.40 | 0.75 | 0.25 | 2.67E-06 | 12 | 50184929 | 50222533 |
| FAM76B | ENSG00000077458 | 0.10 | 1.00 | 0.00 | 2.72E-06 | 11 | 95502117 | 95523573 |
| MED27 | ENSG00000160563 | 0.20 | 1.00 | 0.00 | 2.74E-06 | 9 | 134728315 | 134955254 |
| CPNE3 | ENSG00000085719 | 0.21 | 0.63 | 0.37 | 2.74E-06 | 8 | 87526664 | 87573726 |
| SYT3 | ENSG00000213023 | -0.58 | 0.27 | 0.73 | 2.82E-06 | 19 | 51124564 | 51143138 |
| DCDC2 | ENSG00000146038 | 0.12 | 1.00 | 0.00 | 2.84E-06 | 6 | 24171983 | 24358287 |
| CHCHD3 | ENSG00000106554 | -0.11 | 0.31 | 0.69 | 2.89E-06 | 7 | 132469631 | 132766850 |

Abbreviations: ENSG, Ensembl gene ID; alpha, overall effect of the GreX; w1, contribution weight for African ancestry; w2, contribution weight for European ancestry; chr, chromosome.

**Table S2. Genes associated with white matter hyperintensity using METRO followed by fine-mapping with FOCUS (N=23 genes; P<2.9x10^-6^)**

| **Gene** | **ENSG** | **alpha** | **w1** | **w2** | **P value** | **chr** | **Start** | **End** |
| --- | --- | --- | --- | --- | --- | --- | --- | --- |
| WBP2 | ENSG00000132471 | 1.52 | 0.00 | 1.00 | 7.92E-54 | 17 | 73841780 | 73852588 |
| SH3PXD2A | ENSG00000107957 | 0.00 | 0.50 | 0.50 | 5.78E-20 | 10 | 105353784 | 105615342 |
| DCAKD | ENSG00000172992 | 3.93 | 0.00 | 1.00 | 3.13E-19 | 17 | 43100706 | 43138499 |
| NMT1 | ENSG00000136448 | -0.44 | 0.00 | 1.00 | 1.41E-16 | 17 | 43035360 | 43186384 |
| EFEMP1 | ENSG00000115380 | 0.89 | 1.00 | 0.00 | 1.35E-15 | 2 | 56093102 | 56151274 |
| ICA1L | ENSG00000163596 | -0.27 | 0.36 | 0.64 | 5.44E-12 | 2 | 203637873 | 203736489 |
| NBEAL1 | ENSG00000144426 | 0.57 | 0.00 | 1.00 | 4.91E-11 | 2 | 203879331 | 204091101 |
| WDR12 | ENSG00000138442 | -0.64 | 0.00 | 1.00 | 5.73E-11 | 2 | 203738984 | 203879521 |
| KLHL24 | ENSG00000114796 | -0.99 | 0.00 | 1.00 | 1.01E-10 | 3 | 183353398 | 183402307 |
| NEURL | ENSG00000107954 | 0.93 | 0.44 | 0.56 | 3.28E-10 | 10 | 105253462 | 105352303 |
| CALCRL | ENSG00000064989 | 1.08 | 0.94 | 0.06 | 7.96E-10 | 2 | 188206691 | 188313187 |
| HAAO | ENSG00000162882 | -0.24 | 1.00 | 0.00 | 1.18E-08 | 2 | 42994229 | 43019733 |
| ARMS2 | ENSG00000254636 | -0.62 | 0.00 | 1.00 | 1.64E-08 | 10 | 124214169 | 124216868 |
| GJC1 | ENSG00000182963 | -0.06 | 0.00 | 1.00 | 3.57E-08 | 17 | 42875816 | 42908184 |
| OXER1 | ENSG00000162881 | 0.53 | 1.00 | 0.00 | 7.02E-08 | 2 | 42989639 | 42991275 |
| HTRA1 | ENSG00000166033 | 7.07 | 1.00 | 0.00 | 2.53E-07 | 10 | 124218067 | 124274423 |
| LRRC37A3 | ENSG00000176809 | -0.44 | 0.00 | 1.00 | 2.54E-07 | 17 | 62850248 | 62915598 |
| FBXO31 | ENSG00000103264 | 1.11 | 0.00 | 1.00 | 3.05E-07 | 16 | 87360593 | 87425748 |
| PDCD7 | ENSG00000090470 | -1.81 | 0.00 | 1.00 | 3.54E-07 | 15 | 65409717 | 65426146 |
| CLPX | ENSG00000166855 | 0.40 | 1.00 | 0.00 | 3.54E-07 | 15 | 65440557 | 65477680 |
| EFTUD2 | ENSG00000108883 | 0.00 | 0.50 | 0.50 | 4.49E-07 | 17 | 42927316 | 42976813 |
| UBAP1L | ENSG00000246922 | -1.26 | 0.01 | 0.99 | 5.91E-07 | 15 | 65385098 | 65407538 |
| MAP1LC3B | ENSG00000140941 | 0.37 | 0.13 | 0.87 | 1.94E-06 | 16 | 87417559 | 87438385 |

Abbreviations: ENSG, Ensembl gene ID; alpha, overall effect of the GreX; w1, contribution weight for African ancestry; w2, contribution weight for European ancestry; chr, chromosome.

**Table S3. Genes associated with Alzheimer’s disease (EA GWAS) using METRO followed by fine-mapping with FOCUS (N=69 genes; P<2.9x10^-6^)**

| **Gene** | **ENSG** | **alpha** | **w1** | **w2** | **P value** | **chr** | **Start** | **End** |
| --- | --- | --- | --- | --- | --- | --- | --- | --- |
| CLU | ENSG00000120885 | 0.48 | 0.29 | 0.71 | 4.77E-39 | 8 | 27454434 | 27472217 |
| TOMM40 | ENSG00000130204 | -19.78 | 1.00 | 0.00 | 4.77E-39 | 19 | 45393826 | 45406946 |
| APOE | ENSG00000130203 | 67.11 | 0.28 | 0.72 | 4.77E-39 | 19 | 45409048 | 45412650 |
| APOC4 | ENSG00000224916 | -17.64 | 0.91 | 0.09 | 4.77E-39 | 19 | 45445495 | 45452822 |
| PICALM | ENSG00000073921 | -0.15 | 1.00 | 0.00 | 9.33E-36 | 11 | 85668218 | 85780924 |
| CCDC83 | ENSG00000150676 | 0.28 | 1.00 | 0.00 | 1.01E-26 | 11 | 85566144 | 85631064 |
| CR2 | ENSG00000117322 | 1.26 | 0.00 | 1.00 | 1.87E-25 | 1 | 207627575 | 207663240 |
| MS4A6A | ENSG00000110077 | -0.29 | 0.58 | 0.42 | 1.48E-19 | 11 | 59939488 | 59952139 |
| MS4A2 | ENSG00000149534 | 1.51 | 0.22 | 0.78 | 2.29E-17 | 11 | 59855734 | 59865940 |
| RIN3 | ENSG00000100599 | 3.19 | 1.00 | 0.00 | 7.51E-17 | 14 | 92980125 | 93155339 |
| MEPCE | ENSG00000146834 | -0.22 | 1.00 | 0.00 | 5.62E-16 | 7 | 100025945 | 100031749 |
| PPP1R35 | ENSG00000160813 | 0.31 | 1.00 | 0.00 | 2.40E-15 | 7 | 100032905 | 100034120 |
| CYB561 | ENSG00000008283 | 0.12 | 0.85 | 0.15 | 1.66E-14 | 17 | 61509665 | 61523715 |
| KCNH6 | ENSG00000173826 | -0.27 | 0.62 | 0.38 | 2.33E-14 | 17 | 61600695 | 61626338 |
| USP6 | ENSG00000129204 | -0.38 | 0.98 | 0.02 | 3.47E-14 | 17 | 5019327 | 5078329 |
| TREM2 | ENSG00000095970 | 1.10 | 0.00 | 1.00 | 7.75E-14 | 6 | 41126244 | 41130924 |
| ACE | ENSG00000159640 | 0.65 | 0.24 | 0.76 | 8.53E-14 | 17 | 61554422 | 61575741 |
| ZYX | ENSG00000159840 | 0.17 | 0.00 | 1.00 | 9.46E-14 | 7 | 143078388 | 143088204 |
| FAM131B | ENSG00000159784 | -0.28 | 1.00 | 0.00 | 2.78E-13 | 7 | 143050493 | 143059863 |
| C6orf10 | ENSG00000204296 | -0.39 | 0.00 | 1.00 | 3.04E-13 | 6 | 32256303 | 32339689 |
| BTNL2 | ENSG00000204290 | 0.60 | 0.76 | 0.24 | 7.66E-13 | 6 | 32361116 | 32374958 |
| ATG16L1 | ENSG00000085978 | -0.12 | 0.00 | 1.00 | 1.33E-12 | 2 | 234118697 | 234204320 |
| TREML1 | ENSG00000161911 | -1.07 | 0.96 | 0.04 | 2.31E-12 | 6 | 41117075 | 41122085 |
| ZNF594 | ENSG00000180626 | 0.98 | 0.40 | 0.60 | 2.44E-12 | 17 | 5082830 | 5095163 |
| EPHA1 | ENSG00000146904 | -0.58 | 0.52 | 0.48 | 3.79E-12 | 7 | 143087382 | 143105949 |
| APP | ENSG00000142192 | -0.13 | 0.00 | 1.00 | 1.09E-11 | 21 | 27252861 | 27543446 |
| USP6NL | ENSG00000148429 | 0.08 | 0.40 | 0.60 | 1.33E-10 | 10 | 11502509 | 11653665 |
| SLC39A13 | ENSG00000165915 | -0.12 | 0.00 | 1.00 | 2.95E-10 | 11 | 47428683 | 47438047 |
| PPP4C | ENSG00000149923 | 0.03 | 0.92 | 0.08 | 3.65E-10 | 16 | 30087299 | 30096697 |
| SLC52A1 | ENSG00000132517 | 0.78 | 0.27 | 0.73 | 3.69E-10 | 17 | 4935895 | 4955304 |
| AC008394.1 | ENSG00000233828 | 0.17 | 0.00 | 1.00 | 5.07E-10 | 5 | 86512423 | 86534822 |
| FAM210B | ENSG00000124098 | -0.14 | 0.00 | 1.00 | 9.14E-10 | 20 | 54934030 | 54943719 |
| GCNT7 | ENSG00000124091 | -3.29 | 0.98 | 0.02 | 9.16E-10 | 20 | 55066548 | 55100981 |
| RAPSN | ENSG00000165917 | 0.14 | 0.99 | 0.01 | 1.37E-09 | 11 | 47459315 | 47470695 |
| CD55 | ENSG00000196352 | -0.47 | 1.00 | 0.00 | 1.84E-09 | 1 | 207494864 | 207560149 |
| HLA-DQA2 | ENSG00000237541 | -0.06 | 0.00 | 1.00 | 1.87E-09 | 6 | 32709168 | 32714975 |
| TBX6 | ENSG00000149922 | -0.11 | 0.96 | 0.04 | 1.95E-09 | 16 | 30097114 | 30103245 |
| DYDC2 | ENSG00000133665 | -0.44 | 0.33 | 0.67 | 2.42E-09 | 10 | 82104501 | 82127829 |
| CSTF1 | ENSG00000101138 | -4.15 | 0.00 | 1.00 | 2.62E-09 | 20 | 54967427 | 54981418 |
| CASS4 | ENSG00000087589 | -0.18 | 0.37 | 0.63 | 4.41E-09 | 20 | 54987168 | 55035443 |
| DYDC1 | ENSG00000170788 | 0.30 | 0.39 | 0.61 | 4.65E-09 | 10 | 82095861 | 82116511 |
| LILRA6 | ENSG00000244482 | 0.21 | 0.13 | 0.87 | 5.48E-09 | 19 | 54720737 | 54746649 |
| MME | ENSG00000196549 | -0.23 | 1.00 | 0.00 | 5.99E-09 | 3 | 154741913 | 154901493 |
| NIT1 | ENSG00000158793 | -0.24 | 0.32 | 0.68 | 7.12E-09 | 1 | 161087876 | 161095235 |
| KLF16 | ENSG00000129911 | 0.18 | 1.00 | 0.00 | 1.03E-08 | 19 | 1852398 | 1863578 |
| DEDD | ENSG00000158796 | 0.15 | 0.00 | 1.00 | 1.17E-08 | 1 | 161090764 | 161102478 |
| MAF1 | ENSG00000179632 | -0.11 | 0.00 | 1.00 | 1.52E-08 | 8 | 145159364 | 145162514 |
| TP53INP1 | ENSG00000164938 | -0.05 | 0.66 | 0.34 | 2.05E-08 | 8 | 95938200 | 95961606 |
| KANSL1 | ENSG00000120071 | 0.00 | 0.50 | 0.50 | 3.81E-08 | 17 | 44107282 | 44302755 |
| LACTB | ENSG00000103642 | 0.06 | 0.41 | 0.59 | 4.42E-08 | 15 | 63414032 | 63434260 |
| SHARPIN | ENSG00000179526 | 0.23 | 1.00 | 0.00 | 9.56E-08 | 8 | 145153536 | 145163027 |
| MAPT | ENSG00000186868 | -0.54 | 0.51 | 0.49 | 1.76E-07 | 17 | 43971893 | 44105700 |
| IKZF1 | ENSG00000185811 | -0.13 | 1.00 | 0.00 | 3.33E-07 | 7 | 50343664 | 50472799 |
| CPSF3 | ENSG00000119203 | 0.19 | 1.00 | 0.00 | 3.85E-07 | 2 | 9563780 | 9613230 |
| CCNE2 | ENSG00000175305 | -0.18 | 0.35 | 0.65 | 4.65E-07 | 8 | 95891998 | 95908906 |
| FAM108A1 | ENSG00000129968 | -0.81 | 0.68 | 0.32 | 4.69E-07 | 19 | 1876809 | 1885495 |
| GRN | ENSG00000030582 | 0.18 | 0.84 | 0.16 | 6.31E-07 | 17 | 42422614 | 42430474 |
| ABI3 | ENSG00000108798 | -0.05 | 0.00 | 1.00 | 9.18E-07 | 17 | 47287773 | 47300587 |
| SLTM | ENSG00000137776 | 0.17 | 1.00 | 0.00 | 9.82E-07 | 15 | 59171249 | 59225878 |
| RNF111 | ENSG00000157450 | -0.39 | 0.06 | 0.94 | 1.04E-06 | 15 | 59157374 | 59389618 |
| EPDR1 | ENSG00000086289 | 0.04 | 1.00 | 0.00 | 1.16E-06 | 7 | 37723446 | 37991538 |
| SUPT4H1 | ENSG00000213246 | 0.06 | 0.76 | 0.24 | 1.27E-06 | 17 | 56422536 | 56430454 |
| STYX | ENSG00000198252 | 0.10 | 0.00 | 1.00 | 1.38E-06 | 14 | 53196884 | 53241707 |
| SIGLEC9 | ENSG00000129450 | -0.17 | 0.00 | 1.00 | 1.82E-06 | 19 | 51628163 | 51639908 |
| DDX54 | ENSG00000123064 | 0.22 | 0.36 | 0.64 | 2.02E-06 | 12 | 113594978 | 113623284 |
| TRIB1 | ENSG00000173334 | 0.42 | 0.00 | 1.00 | 2.41E-06 | 8 | 126442600 | 126450645 |
| BLNK | ENSG00000095585 | 0.40 | 0.00 | 1.00 | 2.55E-06 | 10 | 97948927 | 98031344 |
| SLC25A39 | ENSG00000013306 | -0.58 | 1.00 | 0.00 | 2.57E-06 | 17 | 42396993 | 42402238 |
| ICA1L | ENSG00000163596 | -0.06 | 0.41 | 0.59 | 2.68E-06 | 2 | 203637873 | 203736489 |

Abbreviations: EA, European ancestry; ENSG, Ensembl gene ID; alpha, overall effect of the GreX; w1, contribution weight for African ancestry; w2, contribution weight for European ancestry; chr, chromosome.

**Table S4. Genes associated with Alzheimer’s disease (AA GWAS) using METRO followed by fine-mapping with FOCUS (N=2 genes; P<2.9x10^-6^)**

| Gene | ENSG | alpha | w1 | w2 | P value | chr | Start | End |
| --- | --- | --- | --- | --- | --- | --- | --- | --- |
| PVRL2 | ENSG00000130202 | -10.7166 | 0.55221657 | 0.44778343 | 9.67E-81 | 19 | 45349393 | 45392485 |
| TOMM40 | ENSG00000130204 | 10.0181854 | 0.04436227 | 0.95563773 | 1.15E-80 | 19 | 45394477 | 45406935 |

Abbreviations: AA, African ancestry; ENSG, Ensembl gene ID; alpha, overall effect of the GreX; w1, contribution weight for African ancestry; w2, contribution weight for European ancestry; chr, chromosome.
