## Supplementary Tables for "Multi-ancestry transcriptome-wide association studies of cognitive function, white matter hyperintensity, and Alzheimer’s disease"

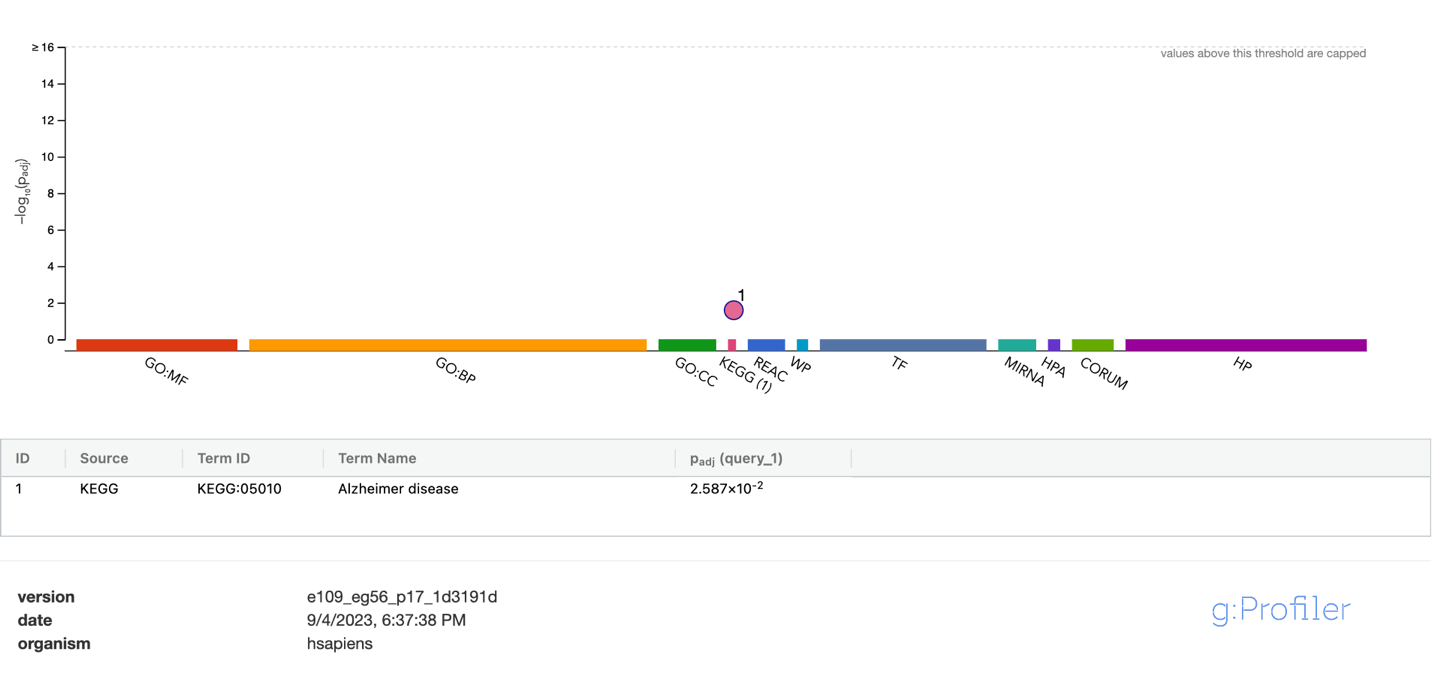

**Figure S1.** **Functional enrichment analysis on the gene set identified by METRO for general cognitive function and AD (N=22 genes; P<2.90x10^-6^).** The top panel consists of a Manhattan plot that illustrates the enrichment analysis results. The x-axis represents functional terms that are grouped and color-coded by data sources, including Gene Ontology (GO): molecular function (MF; red), GO: biological process (BP; orange), GO: cellular component (CC; dark green), Kyoto Encyclopedia of Genes and Genomes (KEGG; pink), Reactome (REAC; dark blue), WikiPathways (WP; turquoise), Transfac (TF; light blue), MiRTarBase (MIRNA; emerald green), Human Protein Atlas (HPA; dark purple), CORUM protein complexes (light green), and Human Phenotype Ontology (HP; violet), in order from left to right. The y-axis shows the adjusted enriched -log_10_ p-values <0.05. Multiple testing correction was performed using g:SCS method (Set Counts and Sizes) that takes into account overlapping terms. The top panel highlights driver GO terms identified using the greedy filtering algorithm in g:Profiler. The light circles represent terms that were not significant after filtering. The circle sizes are in accordance with the corresponding term size (i.e., larger terms have larger circles). The number in parentheses following the source name in the x-axis shows how many significantly enriched terms were from this source.

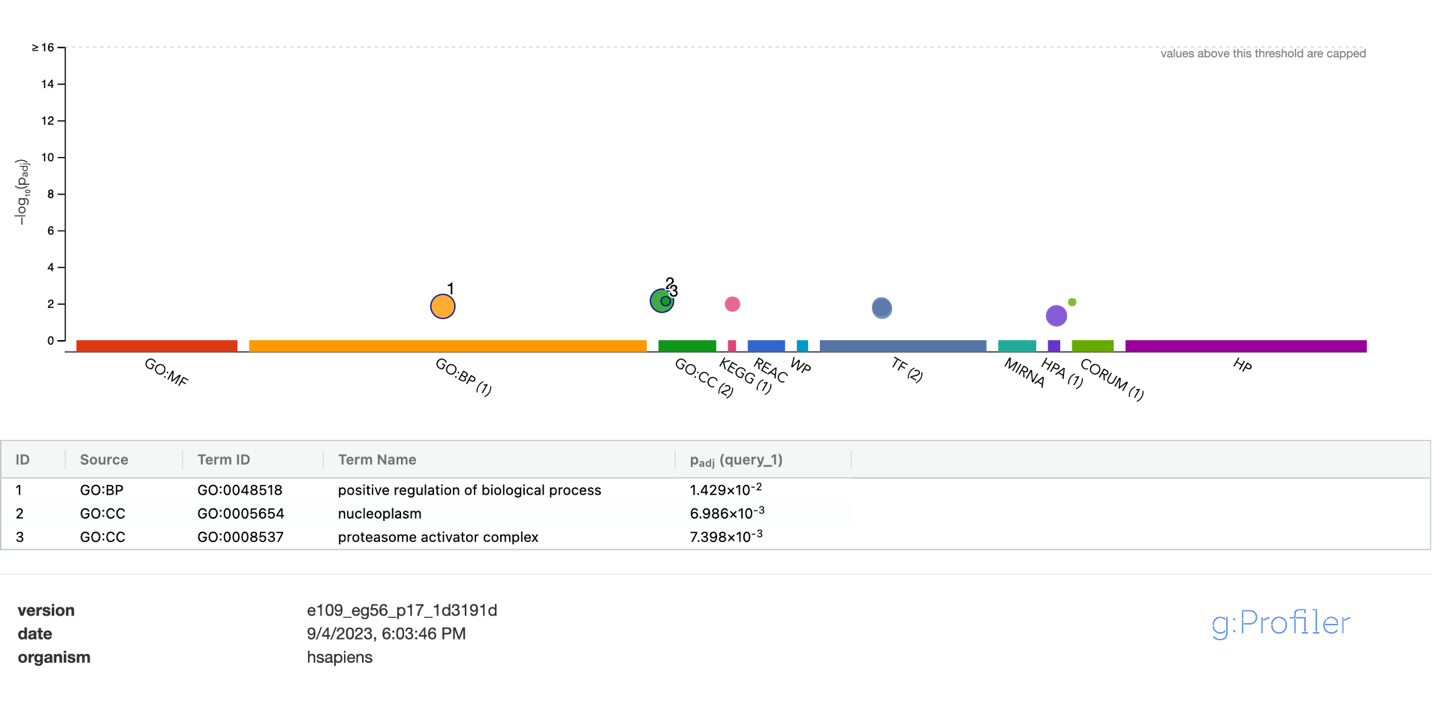

**Figure S2.** **Functional enrichment analysis on the fine-mapped gene set not previously identified by Davies et al. (2018)**(1) **for general cognitive function using METRO TWAS (N=82 genes).** The top panel consists of a Manhattan plot that illustrates the enrichment analysis results. The x-axis represents functional terms that are grouped and color-coded by data sources, including Gene Ontology (GO): molecular function (MF; red), GO: biological process (BP; orange), GO: cellular component (CC; dark green), Kyoto Encyclopedia of Genes and Genomes (KEGG; pink), Reactome (REAC; dark blue), WikiPathways (WP; turquoise), Transfac (TF; light blue), MiRTarBase (MIRNA; emerald green), Human Protein Atlas (HPA; dark purple), CORUM protein complexes (light green), and Human Phenotype Ontology (HP; violet), in order from left to right. The y-axis shows the adjusted enriched -log_10_ p-values <0.05. Multiple testing correction was performed using g:SCS method (Set Counts and Sizes) that takes into account overlapping terms. The top panel highlights driver GO terms identified using the greedy filtering algorithm in g:Profiler. The light circles represent terms that were not significant after filtering. The circle sizes are in accordance with the corresponding term size (i.e., larger terms have larger circles). The number in parentheses following the source name in the x-axis shows how many significantly enriched terms were from this source.

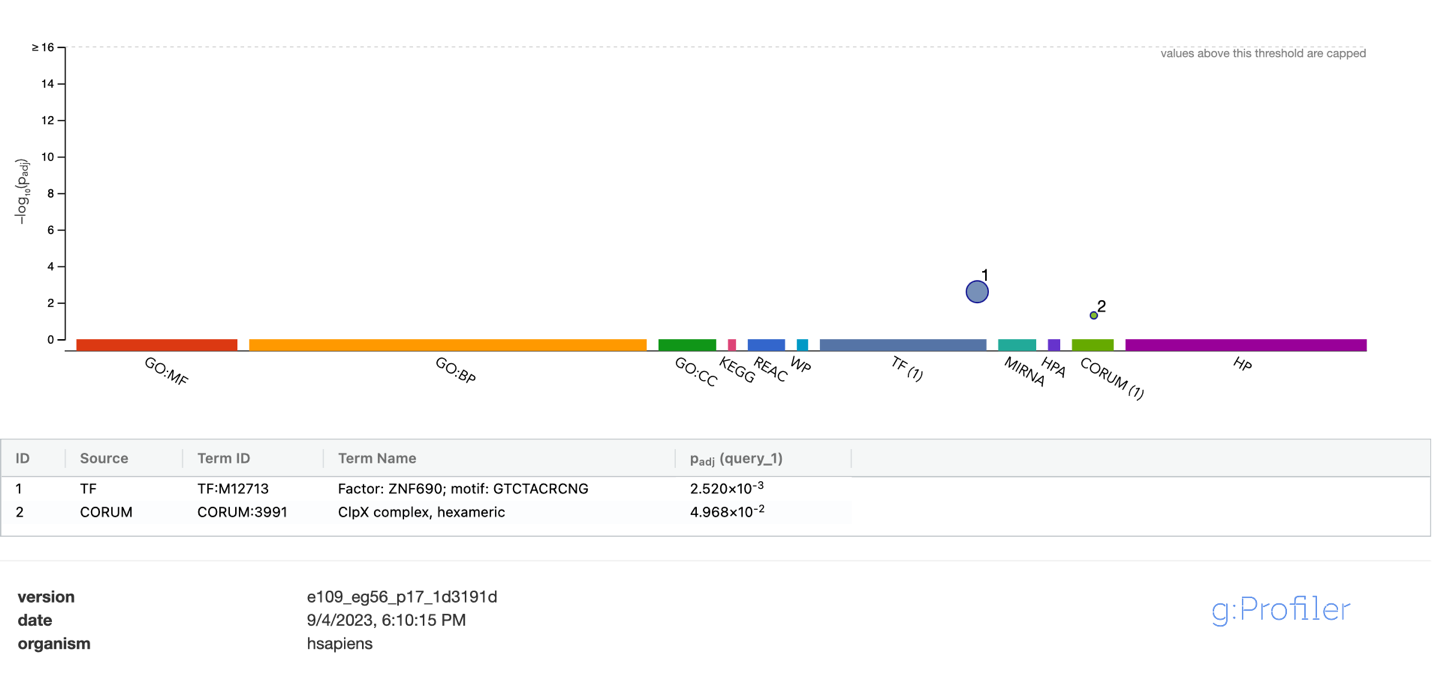

**Figure S3.** **Functional enrichment analysis on the fine-mapped gene set not previously identified by Sargurupremraj et al. (2020)(2) for white matter hyperintensity using METRO TWAS (N=12 genes).** The top panel consists of a Manhattan plot that illustrates the enrichment analysis results. The x-axis represents functional terms that are grouped and color-coded by data sources, including Gene Ontology (GO): molecular function (MF; red), GO: biological process (BP; orange), GO: cellular component (CC; dark green), Kyoto Encyclopedia of Genes and Genomes (KEGG; pink), Reactome (REAC; dark blue), WikiPathways (WP; turquoise), Transfac (TF; light blue), MiRTarBase (MIRNA; emerald green), Human Protein Atlas (HPA; dark purple), CORUM protein complexes (light green), and Human Phenotype Ontology (HP; violet), in order from left to right. The y-axis shows the adjusted enriched -log_10_ p-values <0.05. Multiple testing correction was performed using g:SCS method (Set Counts and Sizes) that takes into account overlapping terms. The top panel highlights driver GO terms identified using the greedy filtering algorithm in g:Profiler. The light circles represent terms that were not significant after filtering. The circle sizes are in accordance with the corresponding term size (i.e., larger terms have larger circles). The number in parentheses following the source name in the x-axis shows how many significantly enriched terms were from this source.

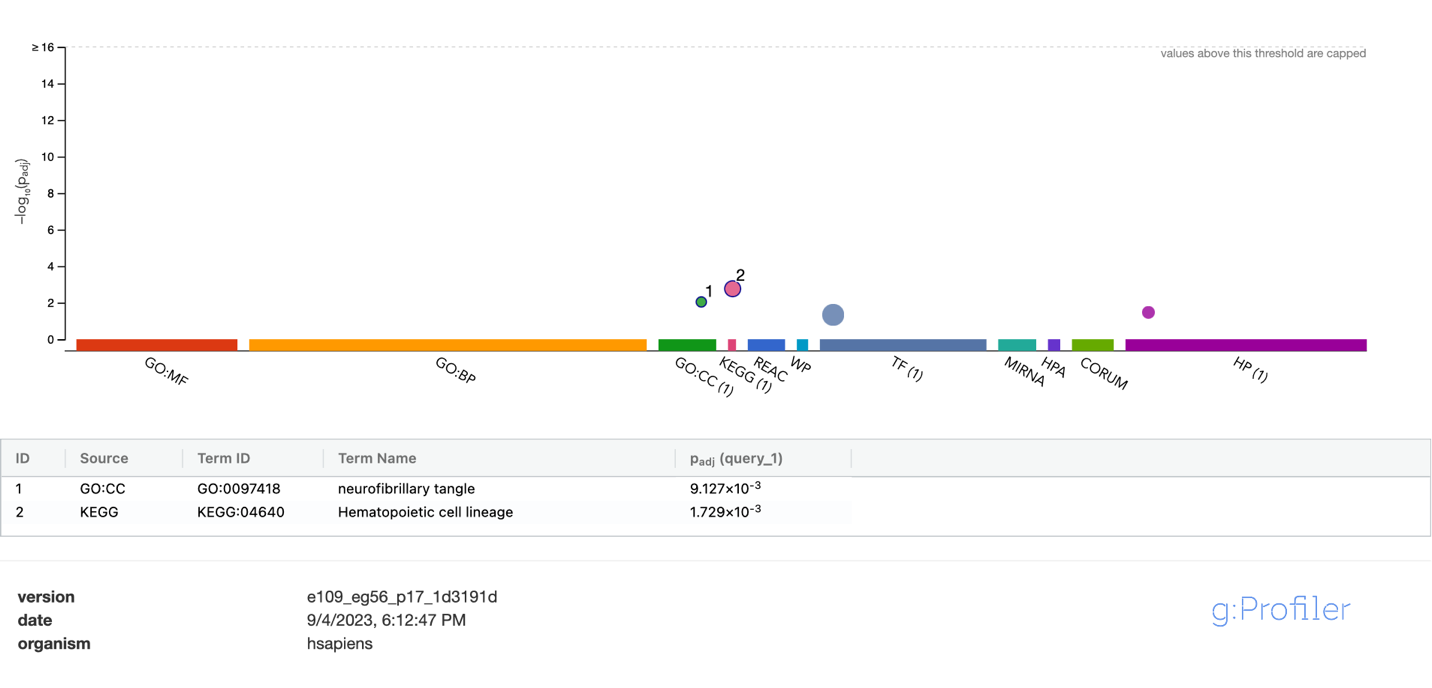

**Figure S4.** **Functional enrichment analysis on the fine-mapped gene set not previously identified by Bellenguez et al. (2022)(3) for Alzheimer’s disease using METRO TWAS (N=45 genes).** The top panel consists of a Manhattan plot that illustrates the enrichment analysis results. The x-axis represents functional terms that are grouped and color-coded by data sources, including Gene Ontology (GO): molecular function (MF; red), GO: biological process (BP; orange), GO: cellular component (CC; dark green), Kyoto Encyclopedia of Genes and Genomes (KEGG; pink), Reactome (REAC; dark blue), WikiPathways (WP; turquoise), Transfac (TF; light blue), MiRTarBase (MIRNA; emerald green), Human Protein Atlas (HPA; dark purple), CORUM protein complexes (light green), and Human Phenotype Ontology (HP; violet), in order from left to right. The y-axis shows the adjusted enriched -log_10_ p-values <0.05. Multiple testing correction was performed using g:SCS method (Set Counts and Sizes) that takes into account overlapping terms. The top panel highlights driver GO terms identified using the greedy filtering algorithm in g:Profiler. The light circles represent terms that were not significant after filtering. The circle sizes are in accordance with the corresponding term size (i.e., larger terms have larger circles). The number in parentheses following the source name in the x-axis shows how many significantly enriched terms were from this source.
